# Global immuno-epidemiology and the persistence of mpox clade IIb in MSM

**DOI:** 10.64898/2026.09.16.26363229

**Authors:** Nils Gubela, Rahel Berghold, Alexander Bartel, Denise Kühnert, Max von Kleist

## Abstract

In 2022, mpox clade IIb spread globally in men who have sex with men (MSM) via sexual contact. By summer 2022, case numbers declined in Europe and Norther America due to behavior change and immunization, with almost no reported cases in 2023, followed by low-level transmission. By autumn 2022, many susceptible individuals at high risk of transmission may have become immunized. Moreover, because individuals with mpox are infectious for only 2–4 weeks, sustaining transmission chains requires considerable infection throughput. This raises the question: How did mpox persist and transition to endemic circulation?

We developed an agent-based model of mpox transmission on the Berlin MSM sexual contact network that incorporates viral shedding kinetics, vaccination, infection-derived immunity, immune waning, and case importations. After calibrating the model, we estimated that the probability of mpox extinction in Berlin exceeded 80% in 2023, when acquired immunity fragmented the transmission network. Extending the analysis to a European MSM metapopulation reduced the extinction probability to less than 60%, whereas incorporating global metapopulation dynamics reduced it to nearly zero. We find that mpox persistence was enabled by epidemic asynchrony: When transmission declined in Europe/the Americas, mpox transmission continued in Asia, long enough for immune waning to partially restore transmission potential in Europe, thereby enabling subsequent re-importation and endemic circulation. These model-based findings are supported by phylogenetic evidence indicating that all active German transmission clusters are either linked to (i) re-importation of clade F from the Americas, or (ii) the emergence of clades C and E linked to Asia.

Our findings on asynchronous transmission across connected global MSM networks highlight the role of global immuno-epidemiology in enabling mpox persistence and emphasize the importance of coordinated international surveillance and control efforts.

## Introduction

Prior to 2022, mpox circulated endemically in Central and West Africa [1], with unknown transmission routes. During 2022, over 85.000 cases were diagnosed globally with most cases in men, and particularly in individuals who identified to have sex with men (MSM), suggesting an important role of sexual contact for mpox transmission [2, 3, 4].

Following the initial epidemic wave, which peaked in summer 2022, reported case numbers declined rapidly in many countries through autumn and winter [5]. This rapid decline was attributed to a combination of factors, including the accumulation of infectionand vaccination-induced immunity, change in sexual behaviour, increased awareness and case detection, and targeted vaccination campaigns [6, 7, 8, 9, 10, 11]. Despite the decline of the initial outbreak in 2022, mpox did not disappear from many affected regions. Instead, several countries experienced continued sporadic transmission following periods of very low incidence, raising questions about the long-term dynamics of the virus outside endemic settings [12]. Persisting low-level transmission is particularly difficult to explain, because mpox is typically infectious only for a short period of time (2-4 weeks) [1, 13, 14], which would require substantial ‘infection-throughput’ to sustain transmission chains. Moreover, by the end of 2022, immunity was acquired in many individuals with the highest risks of infection and transmission.

Recent work suggests that repeated viral introduction may have maintained low levels of transmission after 2022, even when local transmission chains were unlikely to persist [15]. Others speculated that residual local transmission within highly connected sexual networks [16], substantial under-reporting [17], or waning immunity [18, 19, 20, 21] may have contributed. To date, the role of these mechanisms, or their combination in maintaining mpox transmission post-2022 remains poorly understood.

Berlin, Germany, has one of the largest sex-positive MSM populations in Europe and accounted for 44% of all reported mpox cases in Germany [22] and 6% of all reported cases in Europe [23], making it an ideal setting in which to investigate the mechanisms of mpox circulation among MSM. Cases declined in Berlin over the summer of 2022 with only sporadic cases in the winter of 2022/23. No cases were reported for 25 consecutive weeks, until mpox suddenly re-emerged in autumn 2023 in a case without travel history [24]. Subsequently, transmission resumed at low levels, with case numbers gradually increasing through 2024 and 2025. The transition from apparent local elimination to sustained low-level circulation is prototypical across European and American regions and remains poorly understood.

To elucidate these dynamics, we combined prior knowledge on viral shedding kinetics with data-informed sexual networks among MSM in Berlin, as well as information on the vaccination timeline and immune waning, to simulate mpox transmission and the dynamics of sterilizing immunity in Berlin. Based on this integrated model, we subsequently compute the probability that all local infection chains end (local extinction probability). We then consider case importations together with the temporal dynamics of mpox spreading in other European, Asian and American countries to assess global mpox clade IIb persistence, as well as meta-population spreading as a mechanism that may have prevented the extinction of mpox in MSM networks. Finally, we validated our simulations against phylodynamic data, which suggest that exportation and re-importation dynamics, that is, meta-population spread, may have contributed to the persistence of mpox clade IIb among MSM, as illustrated in Fig. 1.

**Figure 1:**
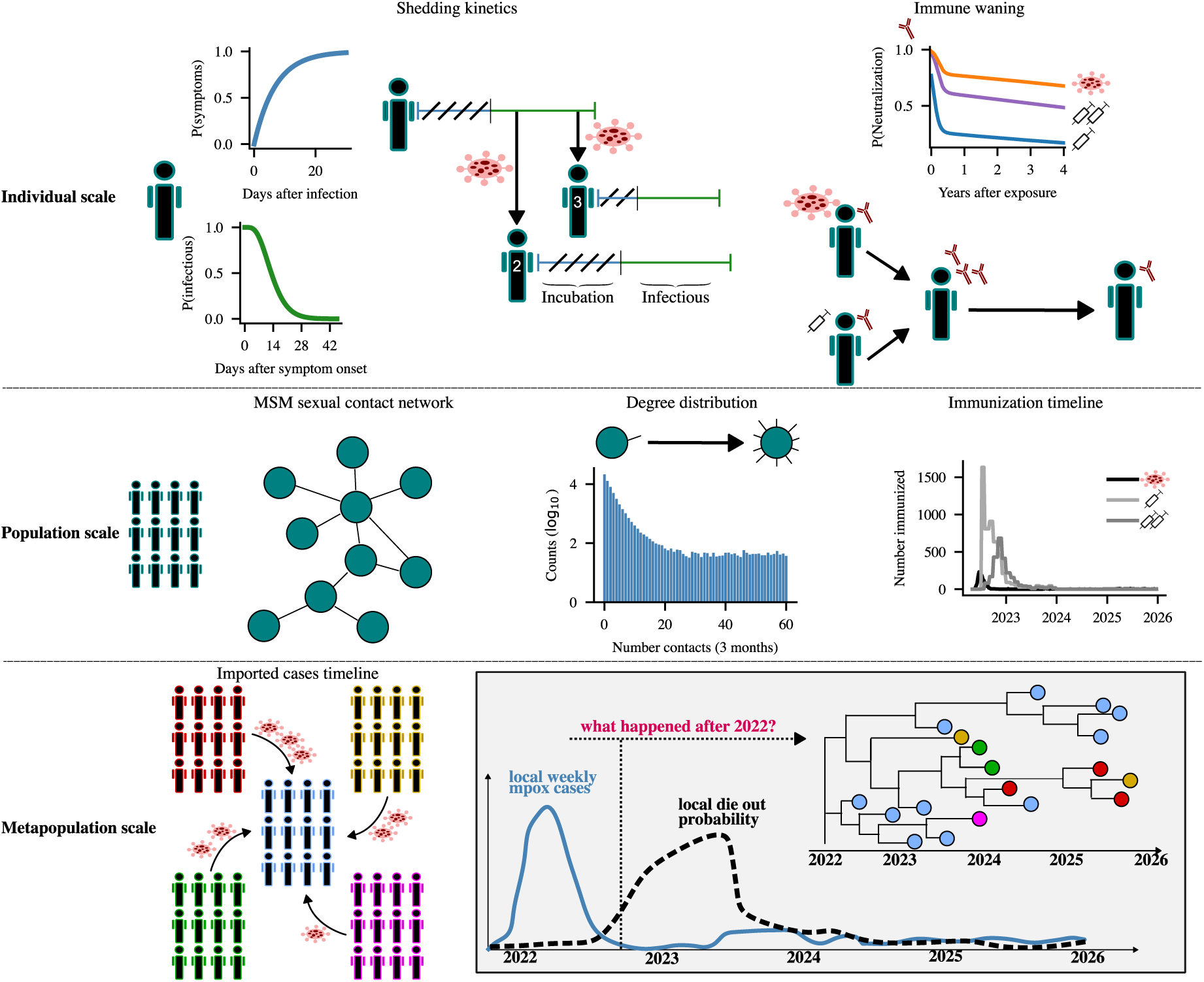
Overview of the data sources and models to estimate local and global extinction probabilities of mpox clade IIb. Top: Individual-level viral shedding kinetics were integrated with vaccine-derived and infection-induced sterilising immunity, including immune waning. Center: Dynamic sexual contact network structure in Berlin MSM, used as a prototypical transmission network, as well as vaccination timeline. Bottom: Temporal incidence patterns in other European, Asian, and American countries, together with case importations, were incorporated to assess extinction probabilities at both local and global scales. Phylodynamic analyses of mpox evolution were further used to evaluate the inferred metapopulation processes underlying the persistence of mpox clade IIb transmission in MSM networks.

## Results

### Model development and calibration

We adapted a previously developed agent-based model of mpox spreading on the Berlin MSM sexual contact network [10, 25], see Fig. 2A. The model takes the size of the Berlin MSM network, the number of sexual contacts, case importations, the vaccination timeline, as well as behavioural changes as input, and was calibrated to the 2022 (May-Oct 2022) outbreak by estimating its two free parameters, the infection rate per sexual mpox exposure *β*, as well as the diagnosis rate *δ* with approximate Bayesian computation using sequential Monte Carlo (ABC-SMC), Fig. 2B. This calibration yielded parameters *β* = 1.7941 week*^−^*^1^ (95% CI: [1.6663, 1.8877]) and *δ* = 0.1036 week*^−^*^1^ (95% CI: [0.1001,0.113]), where the former is within the range of previously reported values [10, 7] and where the latter produces an under-reporting factor of 5.11 (95% CI:[4.77, 5.32]) during the 2022 outbreak, consistent with reported values (5.03; 95% CI:[3.79, 6.57]) derived from risk-adjusted serological studies in Berlin [17, 26]. In order to simulate the succeeding (after October 2022) mpox transmission dynamics in Berlin, we trained a simple model to reflect immune waning dynamics based on available clinical data, see Supplementary Fig. S1. By considering the immune status of individuals at the end of the calibration period (October 2022), as well as immune waning dynamics (compare Supplementary Fig. S2), we continued the simulation until the end of 2025, considering case imports as exogenous infection sources based on Berlin case tracking by the local health authorities. Using this model, simulated cases started to exceed the observed number of cases from spring 2023 onwards, Fig. 2C.

**Figure 2:**
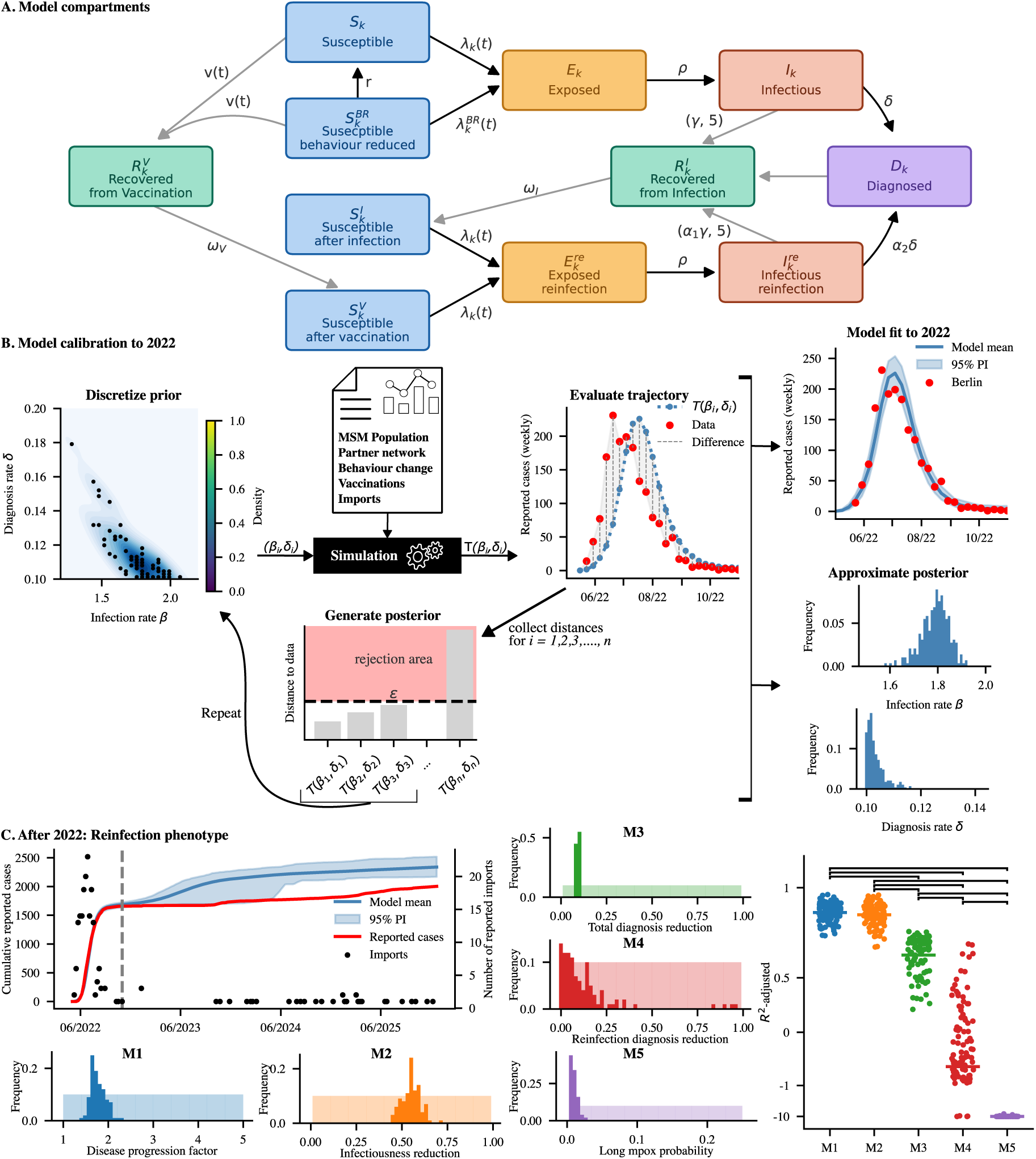
Model compartments and calibration. **(A)** For computational efficiency, the sexual contact network was represented by contact-degree compartments (contact degree *k*), with Markovian (black arrows) and non-Markovian (grey arrows) transitions. Epidemiological dynamics within compartments follow an embedded SIR-IR model (see *Materials & Methods* for details). Background shading of the boxes highlight different classes: Susceptibles (blue), exposed (yellow), infectious (red), recovered (green) and diagnosed (magenta). **(B)** The model was calibrated to the 2022 mpox outbreak in Berlin using ABC-SMC [75], following previous work [10]. **(C)** Left: Cumulative weekly reported cases in Berlin (red line) and imported cases (black dots) vs. a naive model simulation that was only calibrated to the 2022 outbreak (blue line and shading = mean and 95% prediction interval across 292 trajectories). The vertical line marks the end of the 2022 calibration period. Center: Calibration of five competing models (M1-M5) for the post-2022 period. M1: disease duration of breakthrough-and re-infections shorter (blue) or M2: their infectiousness is lowered (orange); M3: lower overall diagnosis probability after 2022; M4: diagnosis probability of breakthrough-and re-infections lower and M5: a proportion of individuals are long-shedders. Show_1_n_7_are the prior-and posterior distributions (shaded areas and solid bars) of the model-specific additional free parameter for the 100 best-fitting trajectories. Right: Model comparison based on goodness of fit for the post-2022 recalibration. Shown are the 100 highest adjusted *R*^2^ values for each model (M1-M5). Connecting lines indicate significant pairwise differences (one-sided Mann–Whitney U test, alternative hypothesis: left model *>* right model) at the *P <* 0.05 level.

The mismatch between model and data prompted us to investigate whether epidemic dynamics may have changed after the initial outbreak. In this context, it was proposed that (a) the relatively high case detection rates during the pandemic phase of 2022 were not maintained after 2022 [27, 17], (b) the phenotype of break-through infections may be distinct from initial infections [28] and that (c) ‘long-mpox’ (individuals with prolonged symptoms and viral shedding kinetics) may have contributed to mpox persistence after 2022 [29]. We therefore added one additional process to the 2022 model respectively, each with one free parameter being calibrated using ABC-SMC (see *Methods section*), see Fig. 2C. Models M1 and M2 share the assumption of a milder presentation of reinfections or vaccination breakthroughs: Model M1 assumed an accelerated progression of breakthrough infections, which indirectly reduces both the potential number of secondary cases emanating from breakthrough infections, as well as diagnosis probability. Model M2 assumed that breakthrough infection is less contagious, because of overall milder presentation, while the duration of the disease is identical to primary infections. Model M3 assumed that the *overall* diagnosis probability of primary and breakthrough infections decreased after the initial outbreak in 2022, while disease progression and contagiousness remained unchanged between breakthrough and primary infection. Model M4 assumed a reduced diagnosis of breakthrough infections and model M5 assumed that a proportion of infected individuals has long mpox, manifested in a prolonged and variable infectious period of 28 to 180 days.

Among these models, M1 and M2 performed significantly better than all other models in terms of adjusted *R*^2^ for the post-2022 period as shown in Fig. 2C (one-sided Mann-Whitney U Test, *P <* 0.05). Simulations with the two best performing models M1 and M2 only differed in the number of actual (diagnosed and undiagnosed) infections occurring after the 2022 outbreak, while the simulated number of diagnosed infections were identical. For M1 (duration of infectiousness) the average number of infections was 1873.68 (95% PI: [1485.33, 2368.48]), and for M2 it was 1562.17 (95% PI: [1113.55, 2061.55]), Supplementary Fig. S3B. In both models, breakthrough infections led to an identical reduction of secondary cases compared to primary infections (on average 0.87 95% PI: 0.82-0.91 transmissions emanating from breakthrough infections, relative to primary infection), see Supplementary Fig. S3D. In M1, the length of the infectious period is shortened by multiplying the recovery rates with a factor of 1.82 (95% CI [1.54,2.15], Fig. 2C, center panel, first graphic). On average, this reduces the infectious period from 2 weeks to 1.1 weeks (95% PI: [0.93, 1.29], compare Supplementary Fig. S3A) and therefore reduces the number of secondary cases emanating from breakthrough infections. In addition, the diagnosis probability for breakthrough infections was reduced in the model as a consequence of accelerated disease progression. The average under-reporting factor over the entire simulation time frame was 5.27 (95% PI: [4.92, 5.52]) and the under-reporting factor for breakthrough infections was 7.42 on average (95% PI: [6.06, 8.79]), see Supplementary Fig. S3C. In M2, infectiousness of breakthrough infections was reduced by a factor of 0.56 (95% CI: [0.47, 0.64]), Fig. 2C. Based on the available data, we cannot distinguish between the two models, as they have an identical impact on the number of reported cases. For convenience, we will report results using M1, while results for M2 are presented in Supplementary Fig. S4-S6. Both models support all conclusions presented herein.

### Return to baseline behaviour and waning immunity partially reconstitute transmission network

In the global North, the 2022 outbreak was contained by transient reductions in behavior, as well as vaccine and infection-induced immunization with variable importance of the individual factors in different countries and hot-spots [30, 7, 8, 31, 10, 11]. All of these factors resulted in a fragmentation of the initial contact network, either by immunization or by reducing contacts (Fig. 3A). We quantify the residual contact network [32], i.e. the contact network excluding individuals who are protected by sterilizing immunity, at four different time points: In the mpox-naive population (05/2022), after the pandemic phase (10/2022), before the reemergence of mpox clade IIb in Berlin (07/2023) [24] and at the end of the simulation time frame (12/2025). In the mpox-naive MSM population, an individual had 4.92 partners in 3 months on average and after the first outbreak (10/2022), an individual had 1.52 susceptible partners in 3 months on average (95% CI:[1.47, 1.59]). Because of immune waning and return to baseline behaviour, the transmission network partially reconstituted in summer 2023 (07/2023) when the average number of sexual contacts with susceptibles increased to 1.71 (95% CI:[1.58, 1.82]), after which it decreased again to 1.52 (95% CI:[1.39, 1.63]) (12/2025), see Fig. 3B.

**Figure 3:**
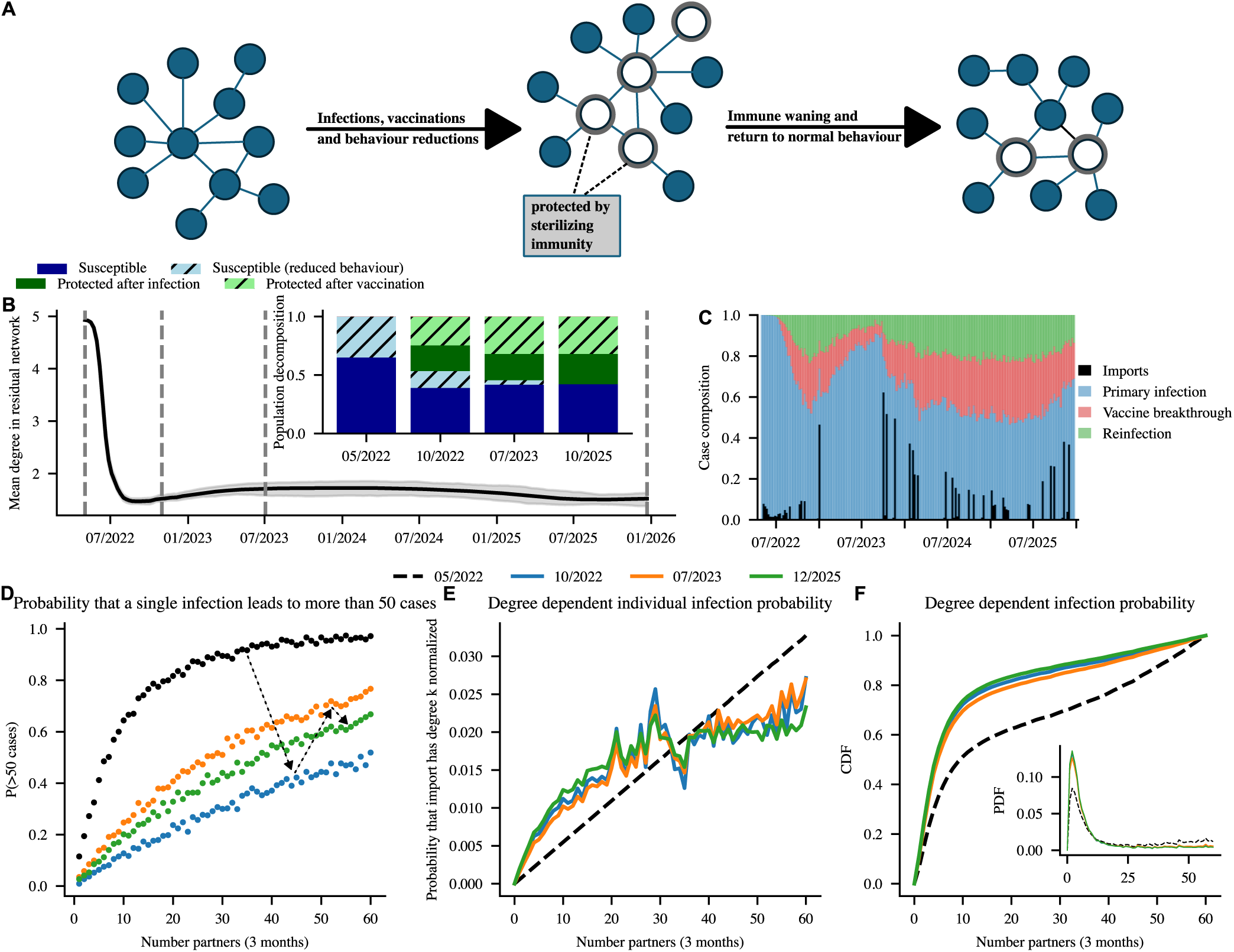
Population immunity gaps. **(A)** Network schematics illustrate connectivity among remaining susceptible individuals (blue nodes) in the network. Infections, vaccination, and behavioural reductions decrease this connectivity (empty nodes), whereas immune waning and return to baseline behaviour restore it and increase the potential for longer transmission chains. **(B)** Mean contact degree of susceptibles over time. Grey shading indicates the 95% CI. Vertical lines mark the start of the simulation (05/2022), the end of the calibration period (10/2022), immediately before mpox re-emergence in Berlin (07/2023), and the end of the simulation (12/2025). The inset shows the population composition by protection and behavioural status at these four time points. **(C)** Weekly case composition by infection type (primary infection, vaccine-breakthrough infection, and reinfection), with imported cases shown separately (black bars). **(D)** Probability that a single infection (e.g. an imported case) of an individual with *k* contacts generates an outbreak with more than 50 secondary cases. For each degree and time point, 1,000 simulations were performed. **(E)** Probability that an infection (e.g. importation) occurs in an individual with *k* contacts, normalised by the size of the corresponding degree class, at the four time points. **(F)** Probability that an imported case has degree *k*. The inset shows the probability density function, and the main panel shows the cumulative distribution function.

We first investigated the proportion of infections attributable to primary infections, vaccine breakthroughs, reinfections, and importations in the Berlin MSM network. According to our simulations, primary infections dominated the 2022 outbreak, whereas the first vaccine breakthroughs and reinfections appeared in autumn 2022 (Fig. 3C). During the phase of very sporadic infections (10/2022–10/2023), a growing fraction were primary infections, after which an equilibrium between primary infections, reinfections and vaccine breakthroughs emerged, Fig 3C. From 2023 onwards, approximately half of all infections were attributable to vaccine breakthroughs and reinfections, while the share of data-informed importations reached its maximum in 2023.

We then analysed the probability that individuals with different numbers of partners can cause an outbreak with more than 50 follow-up cases (Fig. 3D). While the probabilities for the different contact-partner classes increased monotonically with the number of partners, we can see that the outbreak risks are highest in the infection-naive MSM population (05/2022), followed by the population before the re-emergence (07/2023) and the population where we may observe endemic spreading (12/2025). The lowest outbreak risks were observed after the first infection wave (10/2022). Overall, the risk of an outbreak with more than 50 cases was highest when the import occurred in individuals with a large number of contacts.

Therefore, we next investigated how the probability of an exogenous infection entering the network (that is, case importation) depended on the number of sexual partners of the imported case. In the mpox-naive MSM network, the probability that a single individual would import mpox increased linearly with the number of sexual contacts that the individuals has, Fig. 3E, i.e. a person with 40 sexual contacts in the last 3 month had a 20-fold risk of importing mpox in the model, compared with a person with two sexual contacts. For the mpox-experienced MSM networks (10/2022, 07/2023, 12/2025), this relation becomes nonlinear (see Fig. 3E): Since individuals with many contacts are at higher risks to have been exposed to mpox in previous infection waves, a substantial proportion may be protected by sterilizing immunity. Fig. 3F integrates this individual-level information with the abundance of individuals with few vs. many contacts (compare Fig. 1, central panel), showing the overall importation probability in the MSM network. In the infection-naive network, half of all importations occur in individuals with 10 or less sexual partners in the last 3 months, whereas in the mpox-experienced MSM networks (10/2022, 07/2023, 12/2025) half on the importations occur in individuals with 5 or less sexual partners in the last 3 months, because of acquired immunity as shown in Fig. 3A. Thus, exogenous infections become less likely to occur in individuals with many sexual contacts, even though these are the individuals most likely to seed a large outbreak. Instead, imports are more likely to occur in individuals with a low- to mid-range number of sexual partners and these infection chains are more likely to end.

### Evidence for intermittent local extinction in Berlin in 2023

Between February and June 2023, no mpox cases were reported in Berlin (Fig. 2C and Fig. 4A). To further analyse the extent to which endogenous transmission chains may have died out, we quantified the probability that no active mpox infections remained in the MSM network, that is, the probability of local extinction. Foremost, we evaluated different time points. We found that the probability that there was no active mpox transmission chain in the Berlin MSM network peaked in summer 2023, with approximately 80% of simulations having no active cases. This indicates a high likelihood that endogenous mpox transmission within the MSM network had ended by that time (Fig. 4B).

**Figure 4:**
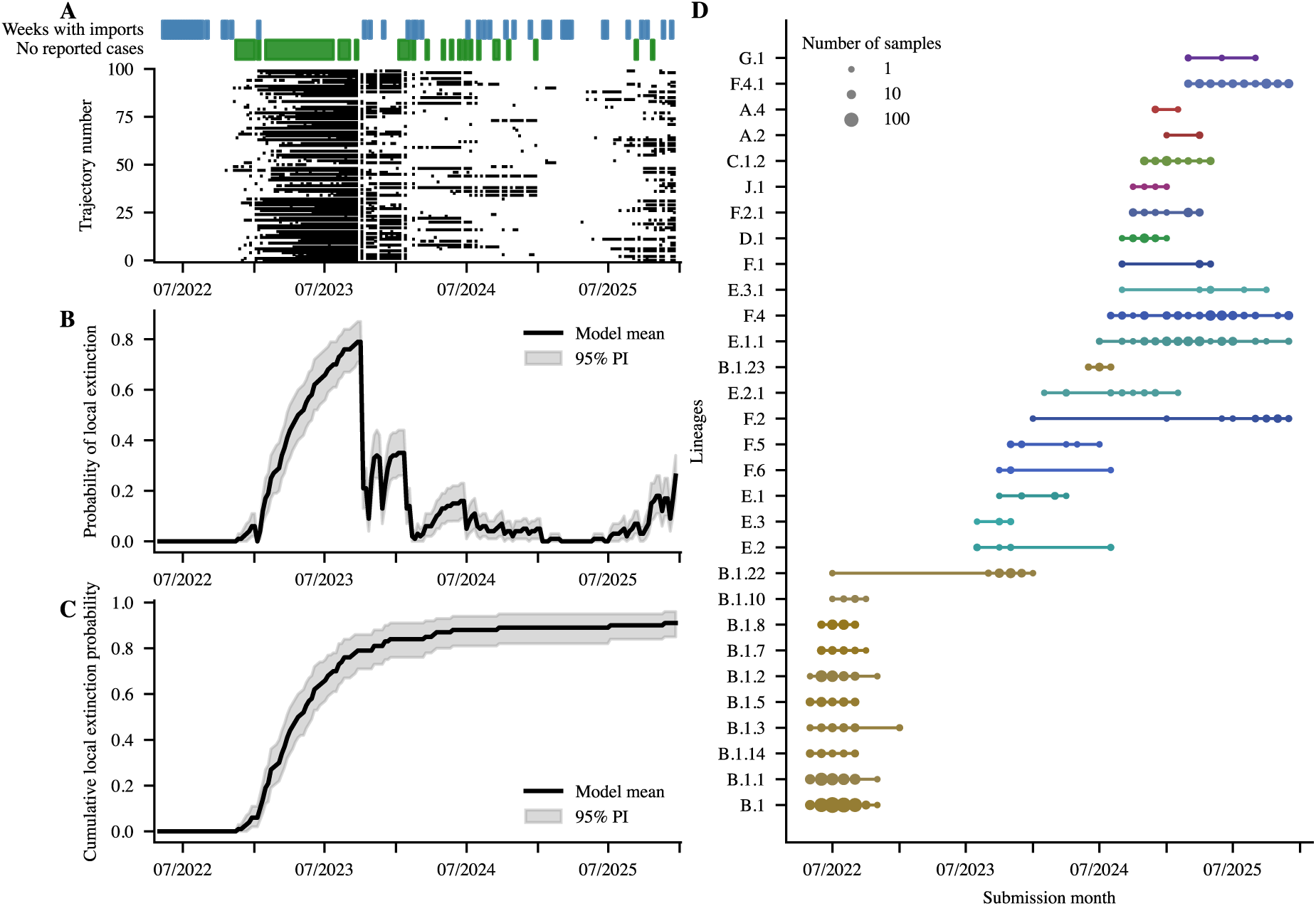
Local mpox clade IIb extinction probability in the Berlin MSM network and sampled lineages. **(A)** Green bars indicate weekly periods with no reported cases in Berlin/Germany and the blue bars indicate weeks with exogenous cases (imported cases). For 100 horizontally plotted stochastic simulations we depicted black brigs if no endogenous infections occurred within a week. **(B)** Weekly probability of no active infections (i.e., extinction) for each model (i.e. vertically counting the frequency of ‘black brigs’ from panel A). Solid lines show the mean across bootstrap replicates and shaded bands the 95% bootstrap confidence interval (CI). **(C)** Cumulative probability of observing at least one week with no active infections (i.e. horizontally counting if a ‘black brigs’ appeared in panel A). **(D)** German mpox Pango lineages by month of sequence submission. Dot size is proportional to the number of sequences, and line segments connect the first and last month in which each lineage was observed.

When following individual mpox simulations longitudinally, we observe at least one time point with no active mpox transmission chain in 91% of all simulations, strongly arguing that endogenous mpox transmission in the Berlin MSM network was very likely to have ended somewhere during summer 2023 or shortly thereafter, Fig. 4C.

To further investigate the nature of mpox transmission during this critical period, we investigated German mpox sequence data (see Fig. 4D). Mpox lineage B.1.22 was the only lineage sampled both before and after 2023, consistent with the possibility of cryptic, unreported circulation. However, this lineage ultimately disappeared in 2024. In contrast, the majority of mpox lineages observed before 2023 are different from the set observed after 2023, which is consistent with our simulations indicating a large probability of local extinction. While case importation may explain why mpox re-emerged in Berlin, other mpox hot-spots from where it could have been imported may constitute similar local extinction probabilities. Thus, the broader question is: Why did mpox clade IIb not go extinct entirely?

### Extinction probability in a desynchronized global metapopulation

The epidemic curves of mpox in several European countries are synchronous between 2022 and 2025 [5]. Reported cases peaked in summer 2022, declined throughout autumn, and reporting largely ceased in early 2023, before cases re-emerged in mid-2023, Fig. 5A. Since previous publications hinted that case reporting may vary substantially across Europe [17], instead of using the absolute number of reported cases, we leveraged information regarding the number of countries reporting mpox to TESSy [33] (see Fig. 5B). We assumed that this number approximates the number of active European mpox hotspots. We then generalized the local mpox extinction probability (compare Fig. 4B, details in *Methods* section) to these hotspots. Using these modelling steps, we computed the probability that mpox goes extinct in all European hotspots simultaneously. These simulations indicated that the probability for mpox to die out in a European MSM meta-population is still considerable (≈ 60%) during 2023 (see Fig. 5C).

**Figure 5:**
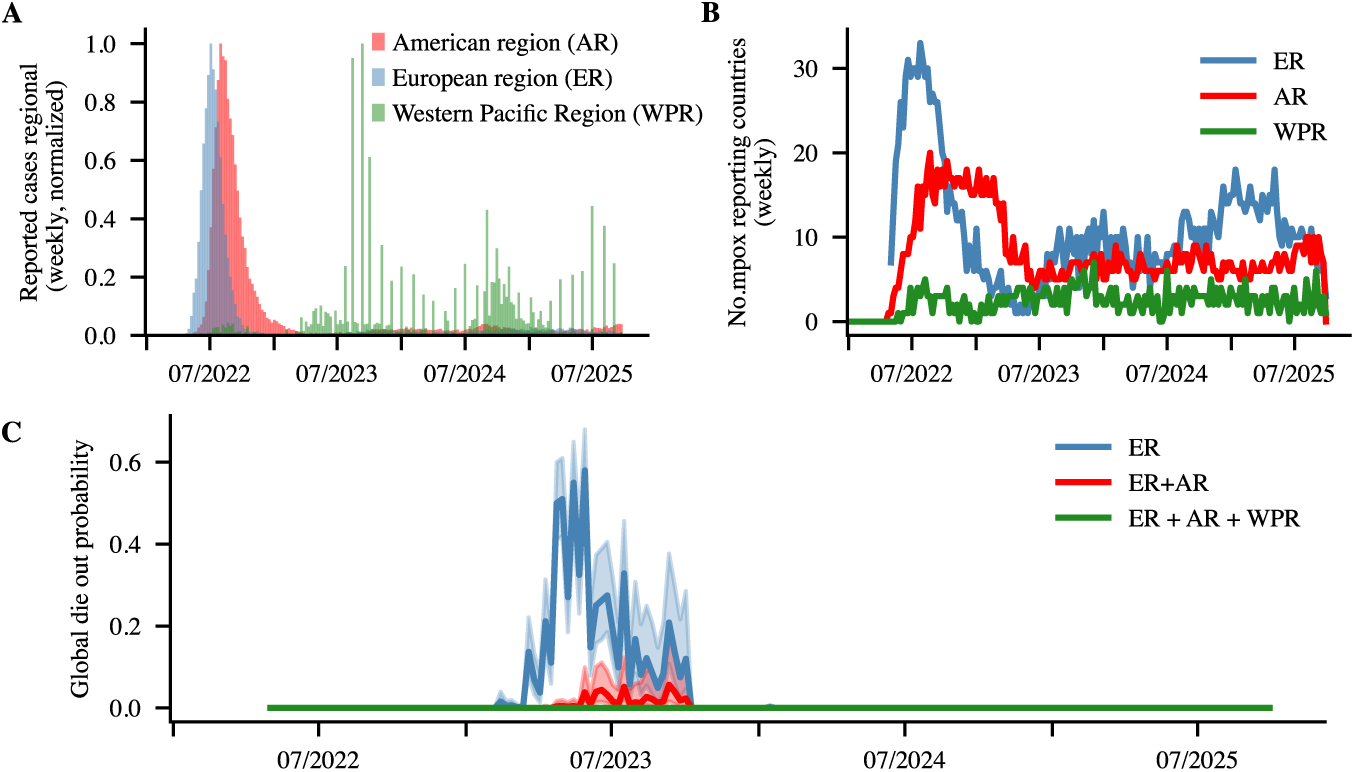
Asynchronous global outbreaks reduce mpox extinction probability. **(A)** Weekly reported mpox cases in the WHO regions: Europe (blue), Americas (red), and Western Pacific (green), normalised to their maximum. **(B)** Number of countries with at least one reported mpox case per week, shown for each WHO region. **(C)** Estimated extinction probability for mpox, calculated for combinations of regions: European region alone (blue); European and American regions (red); and European, American, and Western Pacific regions (green). Estimates are based on Berlin-derived weekly extinction probabilities applied to global surveillance data (see panel B).

A more involved modelling approach based on estimating import rates between European hotspots shows no difference to the approach discussed above as exemplified in Supplementary Methods S1. Next, we added countries from the American region to the metapopulation. The American mpox outbreak was slightly shifted compared to the European epidemic, with more countries reporting cases in the first half of 2023 (Fig. 5A-B). Considering a European-American mpox metapopulation spreading in our simulations still indicated some extinction probability in the second half of 2023 (Fig. 5C).

Lastly, we added countries from the Western Pacific WHO region (including Southeast Asia, China, Japan, Korea, Australia, New Zealand and Oceania) to the analysis. During 2022, these countries reported only sporadic cases associated with travel history or contact with travelers, and no autochthonous mpox transmission was reported [34, 35, 36]. Reported case numbers peaked in the summer of 2023 (see Fig. 5A), approximately one year later than in Europe and the Americas. Including the Western Pacific region with the European and American metapopulation resulted in a vanishing probability of global mpox extinction in MSM, implying that mpox may have been able to persist, because spreading was asynchronous between these three regions.

### Phylodynamic analysis supports asynchronous metapopulation spreading hypothesis

To assess these (bottom-up) findings using a disparate dataset, we collected 9466 mpox sequences from PATHOPLEXUS [37] that were sampled between 2022 and the end of 2025. We then constructed a phylogenetic tree using Nextstrain [38], and analysed phylogenetic clusters containing German sequences after 2023, which may relate to endogenous spreading events. In particular, we were interested in phylogenetic ancestors of these ‘actively circulating clusters’ in 2023. From the phylogenetic tree, we identify circulating transmission clusters belonging to clades C.1.2/E.1.1/E.2/E.3.1 and F.2/F.4 as shown in Fig 6A. In addition, we are interested in the cluster belonging to clade B.1.22/J.1, which appeared before-and after the reporting gap in Berlin. We then constructed BEAST2 phylogenetic trees for the aforementioned clusters. Based on these trees we observe that German transmission clusters within Clades F.2 and F.4 appear to have 2023 ancestry linked to sequences from the US (Fig. 6B), whereas the most recent German sequences from 2025 are inter-dispersed with sequences of US and European origin, see also Supplementary Fig. S7, suggesting an ongoing transatlantic mpox transmission network. A subtree of sequences from clade F.2.1 appears to be linked with Australian sequences from 2024 with no further sequences beyond 2025. Identified circulating German transmission clusters in clades C.1.2/E.1.1/E.2/E.3.1 are phylogenetically related to Asian (Chinese) sequences close to their common root in 2023, from where clades C.1.2/E.1.1/E.2/E.3.1 spawned off, which share a most recent common ancestor with clusters of European (Portuguese) sequences sampled at the end of 2023. These clades sparked distinct outbreaks in Germany in 2024 and 2025. For clade C.1.2, sequence data is largely missing during 2024, suggesting an unobserved intermediary source from where cases may have been imported to Germany before sparking endogenous spreading. More recent German sequences, in particular in clades E.1.1 and E.3.1, are inter-dispersed with European sequences (clade E.1.1) and European and American sequences (E.3.1), indicating several international importation and exportation events, see also Supplementary Fig. S8. Clade B.1.22, from which a single sequence was sampled before the reporting gap in Germany (in 07/2022) re-appeared in Germany at the end of 2023, see Fig. 4D, Fig. 6D and Supplementary Fig. S9. Interestingly, while Fig. 4D may indicate that this clade persisted in Germany, the sequences prior and after the reporting gap are phylogenetically distinct, such that there may be an unknown intermediary infection source that re-ignited autochthonous spreading in Germany in summer 2023 that may have ended at the beginning of 2025.

**Figure 6:**
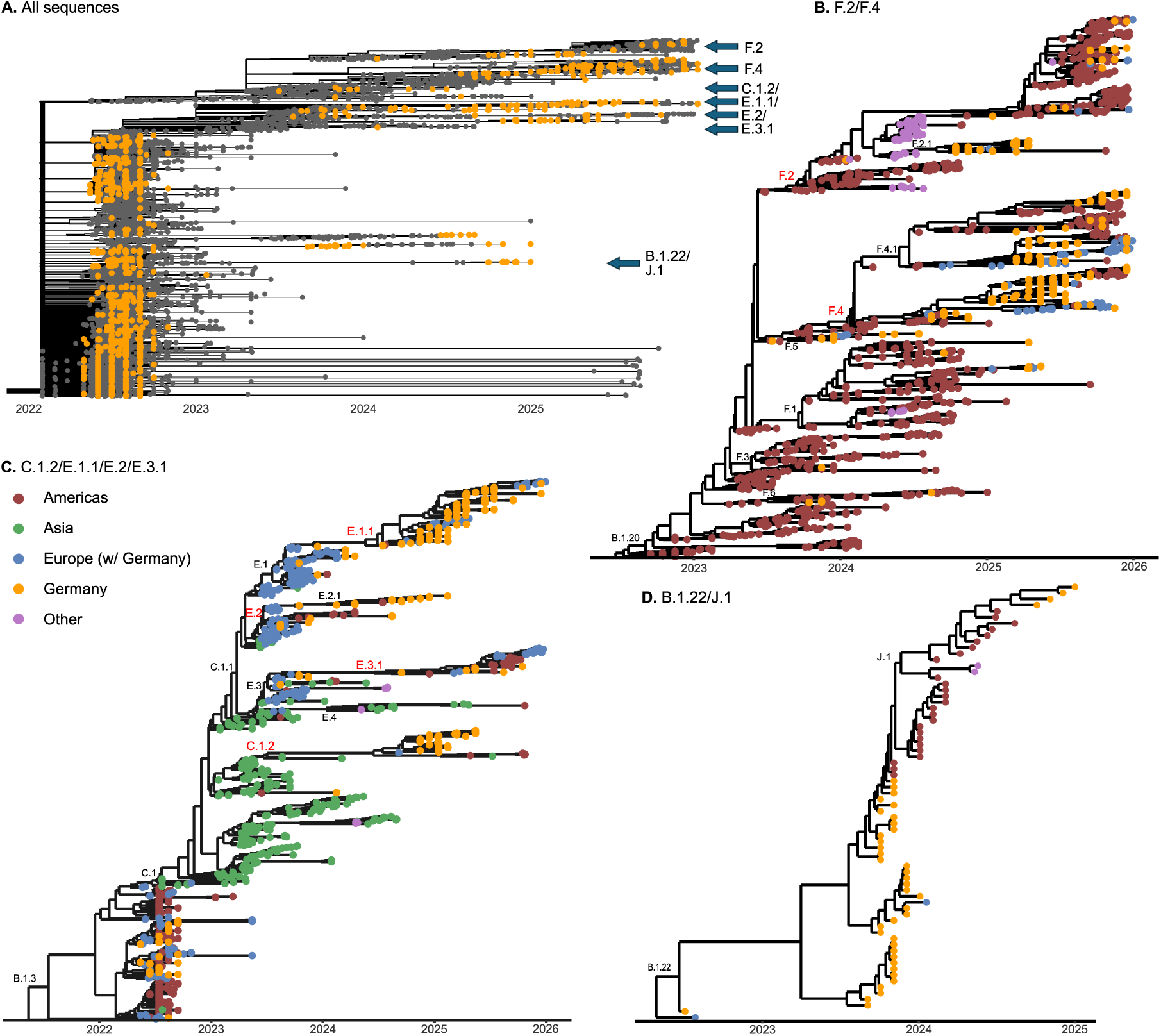
Phylogenetic analysis suggests a 2023 extra-European origin of active German mpox clusters. **(A)** Nextstrain time tree of all available mpox sequences (N = 9466). Active German clusters belonging to subclades F.2/F.4 and C.1.2/E.1.1/E.2/E.3.1 are marked, as well as clade B.1.22/J.1 that appeared before and after the 2023. German sequences are colored yellow. **(B)** BEAST2 timetree of sequences belonging to clades B.1.20, which includes active clusters F.2 and F.4. **(C)** BEAST2 timetree of sequences belonging to clades B.1.3, which includes active clusters C.1.2, E.1.1, E.2 and E.3.1. **(D)** BEAST2 timetree of sequences belonging to clades B.1.22, which includes subclade J.1. Yellow markers: German mpox sequences; grey: non-German sequences; dark red: American sequences; green: Asian sequences; magenta: other origin (Australia/Africa/Oceania)

In summary, the phylogenetic analysis supports dynamics of case re-importation (clades C.1.2/E.1.1/E.2/E.3.1 and F.2/F.4) after 2023 that initiated autochthonous mpox transmission in Germany, from where subsequent exportation of mpox cases (clusters B.1.22) may have occurred. An interesting example are clades F.2 and F.4, which indicate frequent importation-exportation events between Germany and the US, indicative of a transatlantic transmission network, that may partially have been interrupted during the late COVID-19 pandemic.

## Discussion

Some recent studies suggested that mpox has nowadays become an endemic sexually transmitted disease (STD) in MSM [17]. However, in the first half of 2023, only sporadic cases were reported of which most were travel-associated. At the same time, many individuals with frequently changing sex partners had been immunized during the 2022 outbreak and its vaccination campaign. Consequently, 2023 lacked the ‘infection-throughput’ required to sustain autochthonous spreading, as sexual networks of susceptible individuals were likely too fragmented to support prolonged outbreaks following exogenous introductions. This raises the critical question: how did mpox persist among MSM?

To address this question, we developed a multi-scale agent-based model of mpox transmission for the MSM network in Berlin, which we subsequently extended to a global scale. We explicitly considered relevant sexual network structures, which we derived in previous work [10]. Notably, these heavy-tailed networks can have pronounced impact on both the required sterilizing herd immunity beyond which spreading is unlikely, as well as on the burstiness of outbreaks [16, 39, 40].

Using this model, we observe that endogenous circulation of mpox in Berlin is likely to have ended in 2023 (compare Fig. 4B), mainly because acquired immunity fragmented the network of MSM susceptible individuals (compare Fig. 3). Notably, this fragmentation also reduced the probability that exogenous infections (case importations) produce large outbreaks (Fig. 3). Notably, case numbers in Europe and the Americas had similarly declined to very low levels, likely owing to substantial immunization through infection or vaccination [30, 7, 8, 31, 10, 11]. Consequently, if mpox spreading had been globally synchronous, as for example the European epidemic, mpox may have gone extinct in MSM in 2023 (compare Fig. 5C). However, our simulations showed that the global extinction probability was low at any time, Fig. 5C. We predict that the asynchrony of mpox spreading in Europe, the Americas and Asia caused a partial reconstitution of the network of MSM susceptible individuals due to immune waning, which re-enabled endogenous spreading. The outbreak in the Americas was shifted by approximately one month in comparison to Europe, possibly reflecting importation delays and bottlenecks, while the outbreak in Asia was delayed by approximately one year [36]. The latter delay could have been affected by the COVID lockdown, travel restrictions and quarantine requirements, which were only lifted at the beginning of 2023 in China. Notably, there is strong viral phylogenetic evidence supporting re-importation of mpox from Asia and America (Fig. 6) in late 2023: Almost all German cases, and all active transmission chains after 2023 are phylogenetically linked to cases in the Americas and Asia. The initial active German clusters with clades F.2 and F.4 have a clear phylogenetic link to US sequences, whereas more recent sequences from these clades are phylogenetically inter-dispersed, indicating an active transatlantic transmission network. On the other hand, the roots of the active German clusters C.1.2/E.1.1/E.2/E.3.1 in 2023 are most closely related with Asian sequences (mostly from China). Notably, clades C and E have been reported in Japan and South Korea and China in 2023, with possible links to Thailand and other Asian countries [34, 35, 41, 42, 43]. With regards to the C clades there is controversy on the directionality of importation between Asia and Europe. While one phylogeographic study using 76 mpox sequences claims that Portugese C.1.1 sequences, detected in summer 2023, are associated with a Chinese origin [44], another phylogeographic study using 128 mpox sequences inferred the opposite direction of spread, proposing that the Chinese C.1.1 outbreak originated from Portugal [45]. Our analyses do not resolve these competing phylogeographic scenarios, but rather indicate transcontinental meta-population spreading by which asynchronous timing of outbreaks in Asia and Europe may be more consequential than the precise geographic origin of individual lineages. Transmission in Asia may have been seeded by earlier European circulation as indicated in [46], whereas the subsequent re-emergence in Europe may in part be associated with reintroduction of lineages that circulated in Asia at the time.

Our observations regarding meta-population-spreading and the dynamics of the susceptible MSM network have two important immuno-epidemiological implications: First, in contrast to the assertion that immunity to pox viruses may be life-long [47], ‘sterilizing immunity’ to mpox is likely to wane over time (also compare with Supplementary Fig. S1-S2). However, in our model, reinfections had a different phenotype: While we did not explicitly regard symptom severity, we considered re-infections to have a shorter duration-or lesser extent of infectiousness (models M1-2 in Fig. 2C). Findings from others [17] indicate a milder presentation of vaccine breakthrough-and re-infections, which would support this model assumption. From an immuno-epidemiological standpoint it may therefore be important to distinguish between immune-elicited *protection from infection* vs. *protection from disease*, with the former waning faster than the latter.

Second, waning of sterilizing immunity or viral escape from sterilizing immunity have been studied in respiratory viruses that cause more generalized epidemics, such as SARS-CoV-2 and influenza [48, 49, 50]. Respiratory viruses can cause substantial sterilizing immunity in the population, which may limit their transmission potential towards the end of a pandemic wave [48]. In addition to antigenic change, there is evidence in influenza that asynchrony between the northern vs. the southern hemisphere with subsequent re-importation may contribute to maintaining the virus’ transmission potential [51]. Although not a generalized epidemic, we observed that substantial sterilizing immunity to mpox may have been acquired in Europe by the end of 2022, such that the network of mpox susceptible MSM may have been incapable of sustaining endogenous spreading and that because of the asynchrony in spreading dynamics and simultaneous immune waning, mpox did not go extinct (compare Fig. 5). The latter may argue for modelling and monitoring the population immunity landscapes as a public health indicator, in addition to global efforts for pathogen genome surveillance efforts such as the International Pathogen Surveillance Network (IPSN). However, the phylodynamics of lineage B.1.22 are still inconclusive regarding its persistence during the 25 weeks reporting gap in early 2023 [24, 22], which presents itself as a good argument for intensifying pathogen genome surveillance efforts globally.

This work has several limitations: Our model is calibrated to infection dynamics and does not contain relevant information on infection severity. For example, the re-calibration of the model for dynamics post 2022 yielded a shortened duration of the infectious period in breakthrough-and re-infections. We can just assume that a milder presentation of mpox infections (less pustules, or quicker resolution of pustules) is equivalent to a shorted duration of infectiousness in terms of transmission potential. In model M1, a reinfected person has less time to infect others, and an overall lower probability of diagnosis. To disentangle which of these two effects are more important for recapitulating the transmission dynamics, we tested them separately in models M2 (infectiousness reduced) and in M4 (diagnosis probability reduced). M2 performed similarly to M1, while M4 performed significantly worse (Fig. 2C), implying that the reduced infectivity of re-infections is important. Reported mpox reinfections are rare, which can in part be explained by the conditions for diagnosing mpox re-infections: (i) a primary mpox infection must have been diagnosed in the past and (ii) a re-infection shows acute symptoms. The probability of diagnosing the primary infection is already unlikely, given a predicted under-reporting factor of 5 for primary infections in Berlin (which is supported by other studies [52, 10, 17]). Notably, in other European countries under-reporting may be magnitudes larger [17]. Secondly, if re-infections were less symptomatic, there would be less likelihood of symptom-based diagnosis of the re-infection [28]. Conditioned on these biases, a meta-study of reported mpox re-infections found that there was no difference in the clinical presentation between primary-and re-infection, but the duration of infectiousness decreased from approximately 22 days to 13 days [20]. A different study also found that the duration of re-infection is shorter, and reported less mucosal disease in patients with prior infection or vaccination [21]. A milder or faster disease progression after previous infection or vaccination might be due to sustained levels of long-lived memory T-cells targeting conserved MPXV epitopes. The presence of memory T-cells in mpox convalescent donors was previously linked to milder disease [53], but remains an active research topic. On the other hand, long mpox with infectious periods between 33 -141 days [29, 54] had been associated with immunocompromised individuals, such as people living with HIV (PLWH) who have low CD4 counts [29, 55]. We analysed whether long mpox may have been an infection reservoir in Berlin in model M5, which was strongly refuted (Fig. 2C). While HIV prevalence is high in Berlin MSM, the vast majority of cases are virologically suppressed [56] and consequently HIV-related immunodeficiency may have little quantitative impact. In other regions of the world, where HIV prevalence is high and treatment coverage is considerably lower, long mpox may however have more pronounced contribution to infection dynamics and mpox persistence. Another limitation is that we estimated population immunity based on infection and mpox vaccination history. Our analysis did not account for residual immunity from historical smallpox vaccination among individuals older than 50 years or among persons born in countries with longer-lasting smallpox vaccination programmes. Because smallpox vaccination is known to provide cross-protection against mpox [18], which may however only be incomplete [57, 58, 21], and to reduce the risk of severe disease [59], the true level of population immunity may be slightly higher than estimated in this study. At the same time, demographic turnover may further erode immunity within the MSM population at risk. This process is driven by the entry of non-immunised individuals, including younger people at sexual debut and individuals moving to Berlin from regions with little prior mpox exposure and limited vaccine availability. Unfortunately, data on mobility and demographic turnover within MSM populations in Germany are too sparse to parametrise such processes explicitly in our model. As illustrated for clade B.1.22, metapopulation dynamics cannot be resolved in some cases, as viral genetic samples are missing. Likewise, case reporting may also have been incomplete in many regions of the world. In Fig. 5A-B, we try to minimize such biases by utilizing relative estimates (i.e. only considering the number of reporting countries). While this data-normalization may have minimized some sampling biases, it cannot remove them fully.

Finally, at the end of 2025, autochthonous mpox clade Ib transmission occurred in Berlin [60], which nowadays is co-circulating along clade IIb [61]. In this study we solely focus on mpox clade IIb during a time frame before the introduction of mpox clade Ib (2022-2025) for two reasons: relevant transmission parameters, such as the transmission route may be distinct for clade Ib [62, 63]. It is therefore unclear, whether the MSM network utilized herein is appropriate to model clade Ib spreading. Secondly, we focused on the time frame prior to the emergence of mpox clade Ib in Berlin, as there may be cross-immunity between the two virus clades, as well as from from vaccination [64, 65]. While relevant data is too premature at the time of writing, future work may analyze the co-circulation of mpox clade Ib and IIb in MSM, and beyond.

## Materials and Methods

### Case numbers, vaccination data and MSM population

#### Mpox case numbers in Berlin

with a likely MSM route of transmission were provided by the state health authorities (Landesamt für Gesundheit und Soziales (LaGeSo) Berlin), aggregated by either reporting date or date of symptom onset. Case numbers were stratified by the most likely origin (Berlin, outside Berlin, unknown), obtained from case tracking. For fitting, we used the date of reporting, since the model simulates the diagnosis of infected agents rather than the symptom onset. For imported cases, we instead used the date of symptom onset to excite the infectious pressure term *ι*(*t*) (see below), because case imports enter the model as infectious agents. The first cases were reported in calendar week 21 of 2022 (starting May 23), but two patients reported symptom onset already in calendar week 18 (starting May 2). We therefore start simulations in calendar week 18 with two initially infected individuals in our agent based model (ABM) and run the simulation through the end of 2025.

#### Vaccination data

were obtained from the German vaccine monitoring system at the Robert Koch Institute [66]. The total number of vaccinations administered in Berlin was reported monthly. Because the split between first and second doses was available only at the national level, we used the corresponding German proportions to estimate the distribution of first and second doses in Berlin. We approximate weekly vaccination numbers by dividing each month’s total by the number of calendar weeks it contains. For each individual in the ABM who received a first dose, we assign a second dose 2–4 weeks later, drawing from the pool of second doses administered within that time window.

#### Mpox cases in Europe

and the number of countries reporting mpox per week were obtained from the TESSy reporting system [33]. Case reporting time lines for the region of the Americas (North and South America) and the Western Pacific Region (South-East Asia & Oceania) were obtained from the WHO mpox Outbreak dashboard [23].

#### The MSM population

model was adapted from [10], which was sampled from survey data [52]. The survey included variables such as the number of sexual contacts (per three months), MPXV vaccination status, and a binary indicator of whether sexual activity was reduced during the summer of 2022. The survey demographic was divided into vaccinated and non-vaccinated groups. For each subpopulation, a contact distribution was fitted. By early 2023, when the survey was conducted, 18,104 first doses of MVN-BN were administered in Berlin [66]. We sampled 18,104 agents from the vaccinated group and 41,290 agents from the unvaccinated group using the contact distribution, yielding a total Berlin MSM population size in accordance to EMIS [26].

### Agent-based model

We considered four susceptible compartments in the ABM (compare Fig. 2A): susceptible *S*, susceptible with reduced behaviour *S*^BR^, susceptible after vaccination *S*^V^, and susceptible after infection *S*^I^. The infectious compartments are divided into primary infection *I*(*t*) and reinfection *I*^re^(*t*). We denote the number of agents in compartment *C* with *k* expected contacts per week by *C_k_*, the total number of agents with *k* contacts per week by *N_k_*, and the probability that an agent has degree *k* by *P* (*k*). The average degree of the ABM-contact network is then ⟨*k*⟩ = Σ*_k_ kP* (*k*).

In the ABM, infections spread with rate *β* along a susceptible-infected (S–I) connection.

We use the mean-field infectious pressure for the degree-based model [67] to calculate the rate of infection for an individual with contact degree *k*, (*λ_k_*^inf^(*t*)), from the probability that the individual is connected to an infectious agent of degree *k^′^*

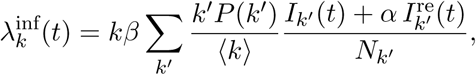

where *α* is the reduction in infectivity for reinfections. A susceptible agent with reduced behaviour is infected at the reduced rate ε*_k_*^inf^(*t*), where *ε <* 1 reflects a lower effective exposure due to behavioural change and was estimated in [10]. The total rate of infection across the population is

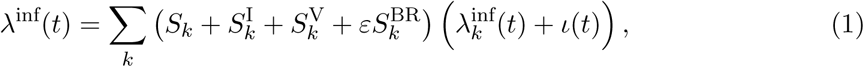

where *ι*(*t*) is an additional hazard accounting for exogenous (imported) infections. We set *ι*(*t*) = 1 whenever an import occurs in the interval [*t, t* + 1), which is large enough to ensure that the average waiting time for an imported infection is less than 10*^−^*^5^. Once the first infection event after time *t* is sampled, it is flagged as an import and *ι*(*t*) is set to 0.

The time between successive infection events is exponentially distributed with rate *λ*^inf^(*t*). Each sampled infection event is assigned to compartment *C* and degree class *k* with probability proportional to *C_k_*(*λ*^inf^(*t*) + *ι*(*t*)).

### Disease progression for primary-and re-infections

After infection, the infected individual progresses to the exposed compartment, in which the agent is not infectious yet. The time from infection to infectiousness was parametrized using clinical data from [6]. Following [10], we set the corresponding transition rate to *ρ* = 1 week*^−^*^1^. Individuals in the infectious compartment are able to pass on the infection, to recover, or become diagnosed. The time to recovery follows an Erlang distribution with rate *γ* = 5*/*2 week*^−^*^1^ and shape *s* = 5 [13, 10]. and diagnosis occurs with rate *δ*, which was estimated from epidemiological data (details below). The transition from the exposed to the infectious compartment was assumed to be identical for reinfections and primary infections, however we considered different recovery and diagnoses rates in reinfections compared to primary infections (*α*_1_*γ* and *α*_2_*δ* respectively). In both cases, the time until recovery is sampled for each individual and logged into an output file in case of diagnosis. Diagnosed individuals are assumed to be non-infectious in the model (i.e., change behaviour accordingly and self-isolate). Individuals eventually transition into the recovered compartment *R*^I^ from both the infectious compartments and the diagnosed compartments.

### Immune waning

Immune waning was modelled by considering both pharmacokinetics and -dynamics: We used a bi-exponential pharmacokinetic model that considered both short-and long-lived neutralizing antibodies *x_s_, x_ℓ_*, which decay at rates *δ_s_*and *δ_ℓ_* [19]. The initial antibody concentration was denoted by *x*_0_, and *f* denoted the fraction of short-lived antibodies. At time *t* after antigen exposure, the expected neutralizing antibody concentrations were then computed by

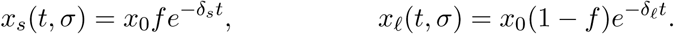

The parameter values *σ* = (*x*_0_*, f, δ_s_, δ_ℓ_*) for one and two doses of MVA-BN were taken from [19] (*δ_l_* = 0.0028*, δ_s_* = 0.23; single dose: *x*_0_ = 80, *f* = 0.89; two doses: *x*_0_ = 673, *f* = 0.94). For infection-derived immunity, we used the same fraction of short-lived antibodies as in the two-dose scheme and set the initial concentration to *x*_0_ = 2000, which is of similar magnitude to reported values for a triple-dose scheme (*x*_0_ = 2211) and therefore represents a conservative assumption for the waning time of infection-induced immunity [68].

The neutralisation probability at antibody concentration *c*(*t, σ*) = *x_s_*(*t, σ*) + *x_ℓ_*(*t, σ*) was then computed using classical pharmacodynamic relationships [48]:

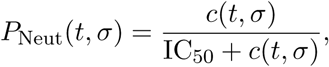

where IC_50_ is the concentration at which 50% protection from mpox infection is achieved. We estimated IC_50_ by choosing the value that minimised the discrepancy between the model-predicted neutralisation probability and clinical vaccine effectiveness estimates measured two weeks after the first dose, VE(14, 1-dose) [69, 70, 71, 72, 73, 74], and two weeks after the second dose, VE(14, 2-dose) [70, 71, 72, 73]:

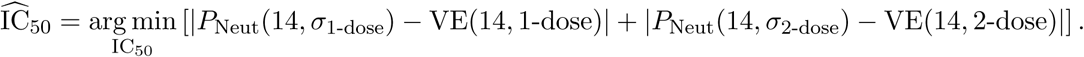

### Model calibration

Model calibration was performed based on Approximate Bayesian Computing based on Sequential Monte Carlo (ABC-SMC) [75] in two steps. First, we calibrated the two free parameters of the ABM to the 2022 outbreak data. Subsequently, we assessed competing hypotheses which may be able to explain the epidemic dynamics post-2022. Each of these competing hypotheses required the estimation of one free parameter.

#### Calibration to the 2022 outbreak

The basic ABM model has two free parameters: The infection rate *β* and the diagnosis rate *δ*. We performed three steps of ABC-SMC on the pandemic phase between May and October 2022. In the first round, priors were *β* ∼ *U* (0, 4) and *δ* ∼ *U* (0.1, 0.26), with the bounds on *δ* corresponding to a diagnosis probability of 18–40% [52]. Each prior was evaluated at 100 equidistant points, and one trajectory was sampled per unique parameter pair (*β, δ*), yielding 10,000 trajectories in total. Trajectories *T* (*β, δ*) were scored against the data *D* using two Euclidean distances: *d*(*T* (*β, δ*)*, D*) on the daily reported case values, and *d*(*C*[*T* (*β, δ*)]*, C*[*D*]) on the cumulative values, where *C*[*X*] = { *_s<t_ X_s_*}*_t_* denote the cumulative number of reported cases of a time series *X*. Trajectories with a distance of 250 or less and a cumulative distance of 750 or less were retained as the posterior sample. In the second round, 10,000 new parameter pairs were drawn from a Gaussian kernel fitted to this posterior, and trajectories within a distance of 150 and a cumulative distance of 400 were retained. In the third round, 100,000 parameter pairs were drawn from a Gaussian kernel fitted to the second-round posterior, and trajectories within a distance of 150 and a cumulative distance of 250 were retained, yielding a final set of 292 (*β, δ*) pairs, each paired with a fixed random seed for reproducibility.

#### Model calibration for the post-pandemic phase

To reproduce the mpox epidemic dynamics post-2022, we calibrated and compared five competing models. Each model has one additional free parameter, which we fit by line search over 100 candidate values for each of the 292 selected (*β, δ*) pairs. For each model, we retained the 100 trajectories over the full time frame with the smallest cumulative distance to the data. In the baseline model, the scaling factors *α*_1_, *α*_2_, and *α* are all fixed to 1 (i.e., reinfections behave identically to primary infections in duration, diagnosis rate, and infectivity). The parameters (*β, δ*) are taken from the calibration to the pandemic phase (year 2022). Each of the five models below is calibrated based on one additional free parameter, while keeping the values of (*β, δ*) derived in the previous step:

- **Reinfection duration scaling (M1):** scales the transition rate through the reinfection compartment, *α*_1_*γ*, where *γ* is the baseline transition rate (see Fig. 2A) and *α*_1_ ∼ *U* (1, 5).
- **Reinfection infectivity scaling (M2):** scales the infectivity of reinfected individuals via the parameter *α* ∼ *U* (0, 1) from (1).
- **Reinfection diagnosis scaling (M3):** scales the rate at which a reinfection is diagnosed, *α*_2_*δ*, with *α*_2_ ∼ *U* (0, 1).
- **Diagnosis decrease after October 2022 (M4):** a new diagnosis rate *δ*_2_ = *α*_3_*δ*, with *α*_3_ ∼ *U* (0, 1) is fitted after October 2022, reflecting a uniform decrease in diagnosis risk following the peak of the outbreak.
- **Long-mpox (M5):** Each infection has a probability *p*_LM_ ∼ *U* (0, 1) of becoming a ”long spreader,” in which case its infectious period is drawn uniformly at random from (28, 180) days [29] instead of the baseline duration. The free parameter fitted for this model is *p*_LM_.

### Phylodynamic analysis

The phylodynamic analysis was performed in two steps. First, all available clade IIb MPOX sequences sampled between 2022 and the end of 2025 were downloaded from PATHOPLEXUS https://pathoplexus.org/ [37] (reference list attached in Suppl. Table ST1). A timed Maximum-likelihood tree of all mpox sequences was built using the Nextstrain MPOX workflow for clade IIb [38] [76]. Some adjustments were made to the filtering to include all sequences which passed quality filtering, which yielded 9466 sequences from 36 countries. Filtering was performed as part of the Nextstrain workflow, which removes sequences below a minimum length of 100,000, as well as those with an excess of private mutations relative to their neighbors in the Nextclade [77] reference tree and additionally excludes a curated list of sequences flagged as duplicates, recombinants, or otherwise problematic. The pipeline additionally masks parts of the genome from the alignment: the first 1350 and last 6422 base pairs, along with several repetitive regions. From the nextstrain tree, clusters of interest (clusters that contained recent German mpox sequences = ‘circulating clusters’; and clusters that contained German mpox sequences before and after 2023 = potentially ‘persistent endogenous clusters’) were chosen for subsequent analysis. We excluded two sequences which showed branches substantially longer than those of all other sequences in the substitution tree (PP 006WPPZ, PP 006WOQX), as well as those for which only the sampling year was known. If only the sampling month of a tip was present in the data, the 15th of that month was taken as tip date. Each cluster comprised a lineage and all of its descendants: ‘circulating clusters’ B.1.20 with F.1–F.6 (1500 sequences) and B.1.3 with C.1 and E.1–E.4 (951 sequences), as well as potentially ‘persistent clusters’ B.1.22 with J.1 (72 sequences). Trees of those clusters were created using BEAST2 version 2.7.7 with a constant coalescent. We used a strict clock with a HKY substitution model and estimated frequencies with gamma rate heterogeneity using 4 categories. Two MCMC chains were run for each cluster with convergence requirements leading to chain lengths of 255M for B.1.20, 200M for B.1.3, 100M for B.1.22 and a pre-burnin of 10M states each. Convergence was inspected using Tracer version 1.7.2 and it was ensured that all ESS were ≥200. Burn-in was determined per cluster by visual inspection of the trace files in Tracer and discarded before further analysis: 11% for B.1.20, 50% for B.1.3, and 10% for B.1.22. After agreement of the two replicates for each cluster was verified, MCC trees were created using TreeAnnotator v.2.7.7 with mean heights. Plotting was performed using ggtree. Lineage assignments for individual sequences were taken from the Nextstrain mpox ingest workflow, which runs Nextclade to place each sequence on a curated reference tree and assigns it the lineage of its nearest node. These tip assignments were then mapped onto the BEAST2 trees by labelling each internal node with a lineage if all of its descendants belonged to that lineage or one of its sublineages. Parameter distribution priors for the BEAST2 analysis are given in Supplementary Table ST1.

## Acknowledgments

We thank all participating laboratories for making mpox sequencing data publicly available. This work has been partially funded by the Federal Ministry of Research, Technology and Space (BMFTR) project EPISERVE (funding ID: 031L0324C), member of the German Modeling network for severe infectious diseases (MONID). Funded by the Deutsche Forschungsgemeinschaft (DFG, German Research Foundation) under Germany’s Excellence Strategy – The Berlin Mathematics Research Center MATH+ (EXC-2046/1, project ID: 390685689). The authors would like to thank the HPC Service of FUB-IT, Freie Universität Berlin, for computing time [78].

## Data availability

The datasets generated during the current study, as well as all input parameters for the simulations, are available via GitHub at https://github.com/KleistLab/MPoxAfter2022/tree/main/results and https://github.com/KleistLab/MPoxAfter2022/tree/main/inputs, respectively. Sequence data was downloaded from PATHOPLEXUS as described above. BEAST2 control files are available at https://github.com/KleistLab/MPoxAfter2022/Phylogeny. All manuscript figures can be reproduced using https://github.com/KleistLab/MPoxAfter2022/blob/main/notebooks/figures.ipynb.

## Code availability

Codes are available via GitHub at https://github.com/KleistLab/MPoxAfter2022 and via Zenodo https://doi.org/10.5281/zenodo.22765868.

## S1 Supplementary Methods: Simulation of Metapopulation Spreading

To explore the extinction mechanism in a simplified setting, we simulated a hierarchical country network. We assumed that importations into country *i* occur with rate *w_i_λ*(*t*), where *w_i_* is a weight proportional to the size of the MSM population in country *i* and *λ*(*t*) is a time-varying importation rate shared across countries. The state of country *i* is denoted by *A_i_*(*t*) and is given by 1 if the mpox outbreak is active in *i* and 0 otherwise. Conditional on being inactive at the previous time step, the probability that country *i* becomes active due to importation is

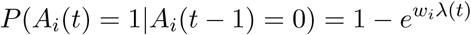

The importation rate *λ*(*t*) was parametrized using the number of reporting countries, together with a linear time term and an intercept, and fitted to the Berlin importations using Poisson regression, yielding

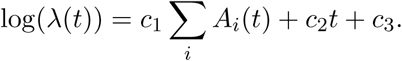

At each time step, the country-level extinction probability was taken directly from the Berlin simulations. The resulting simulated timeline of the number of reporting countries closely resembles the empirical pattern. The inferred global die-out probability is again approximately zero for most time points, with a pronounced spike in spring and summer 2023 and an elevated risk of global die-out persisting until summer 2024, see Figure S10.

---

**Supplementary Figure S1:**
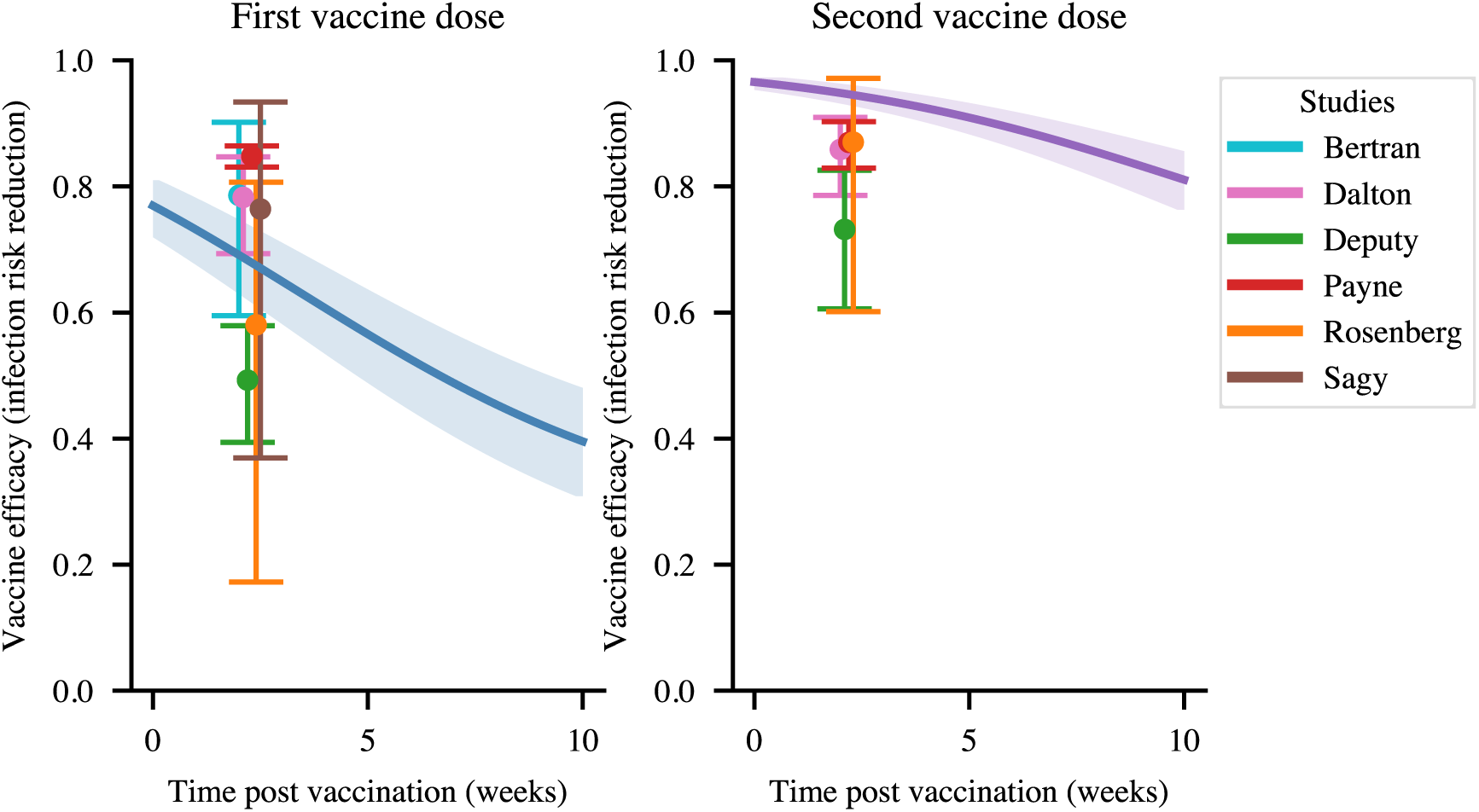
Estimation of vaccine potency IC_50_. The vaccine antibody kinetics are translated to vaccine efficacy (infection prevention) with an emax function with one free parameter IC_50_. This is fitted by minimizing the distance of the 1-dose and 2-dose pharmacodynamics to clinical data two weeks after the first dose [69, 70, 71, 72, 73, 74] and two weeks after the second dose [70, 71, 72, 73]. Shaded areas highlight the 95% CI computed using the pharmacokinetic parameter ranges reported in [19].

**Supplementary Figure S2:**
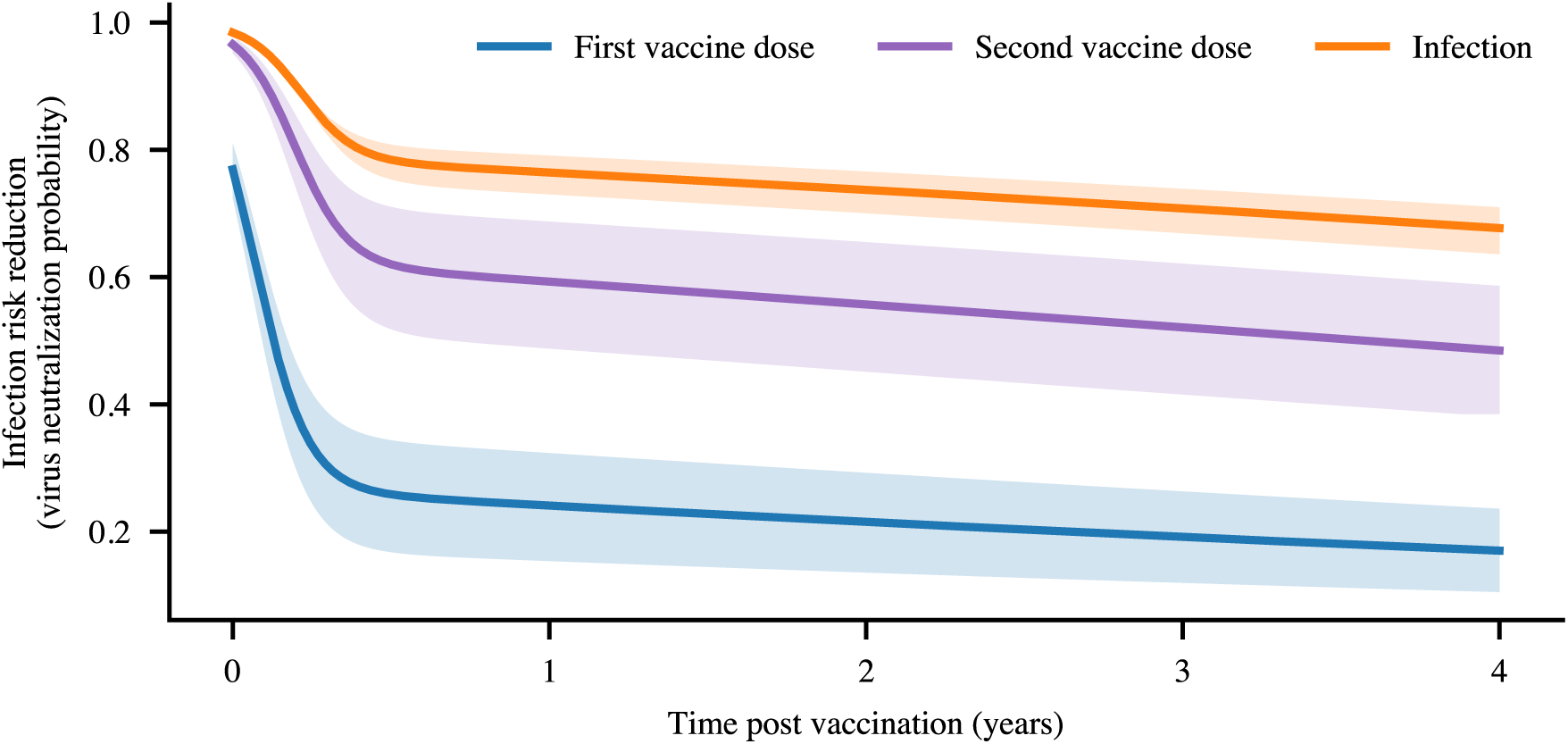
Simulated efficacy of 1-dose and 2-dose vaccination against infection, as well as infection-induced immunization. Blue: first vaccine shot; magenta: second vaccine shot; orange: infection-acquired immunity. Shaded areas highlight the 95% CI computed using the pharmacokinetic parameter ranges reported in [19].

**Supplementary Figure S3:**
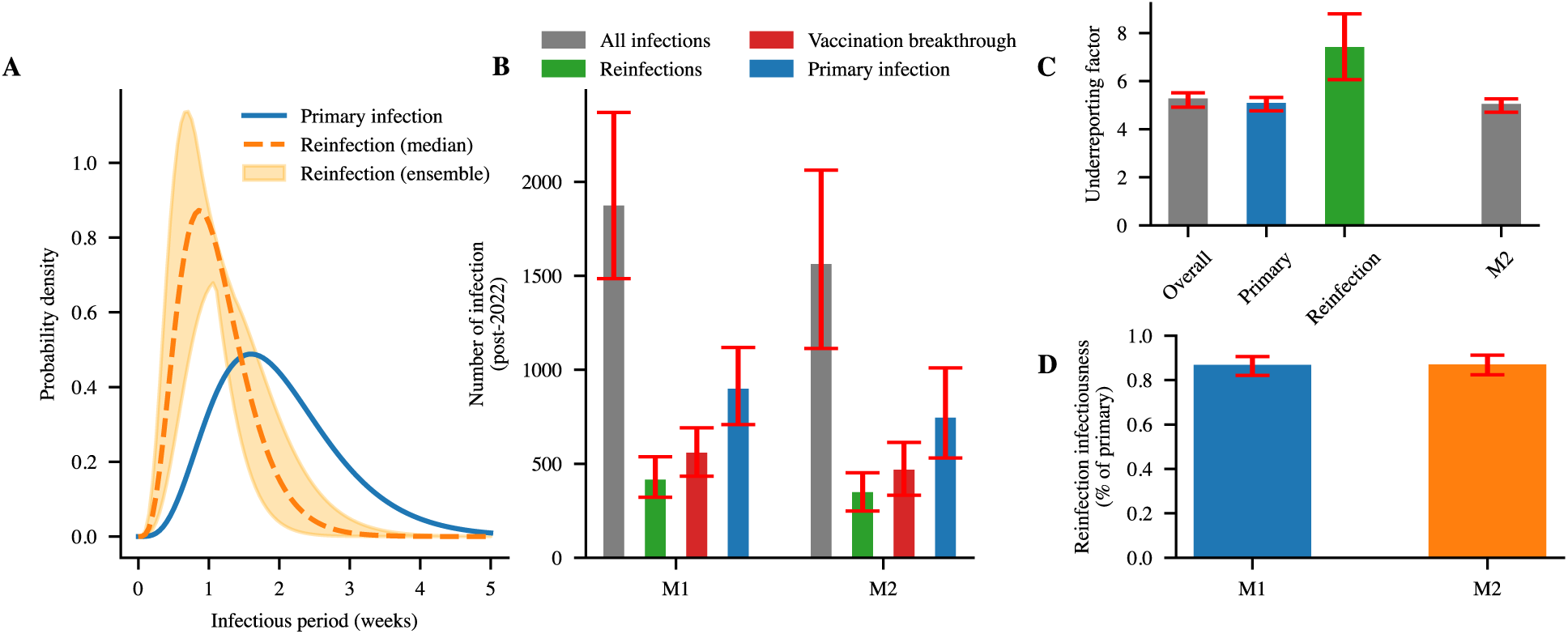
Infection durations, incidence composition, and inferred reporting. **(A)** Distribution of infectious-period lengths for primary infections (blue) compared with reinfections in model M1. The shaded region shows the ensemble of inferred probability density functions; the dotted line indicates the median density at each discrete duration. **(B)** Post-calibration infections (after October 2022) in the two best-performing models (M1 and M2). Bars show mean counts; grey denotes total infections, with primary infections, reinfections, and vaccine-breakthrough infections shown in blue, green, and red, respectively. Error bars indicate the 95% probability interval from 100 simulations. **(C)** Time-varying under-reporting factor over the full simulation period for models M1 and M2. For model M1, under-reporting is shown separately for primary infections and reinfections. **(D)** Relative infectiousness of reinfections compared with primary infections in model M1 and M2.

**Supplementary Figure S4:**
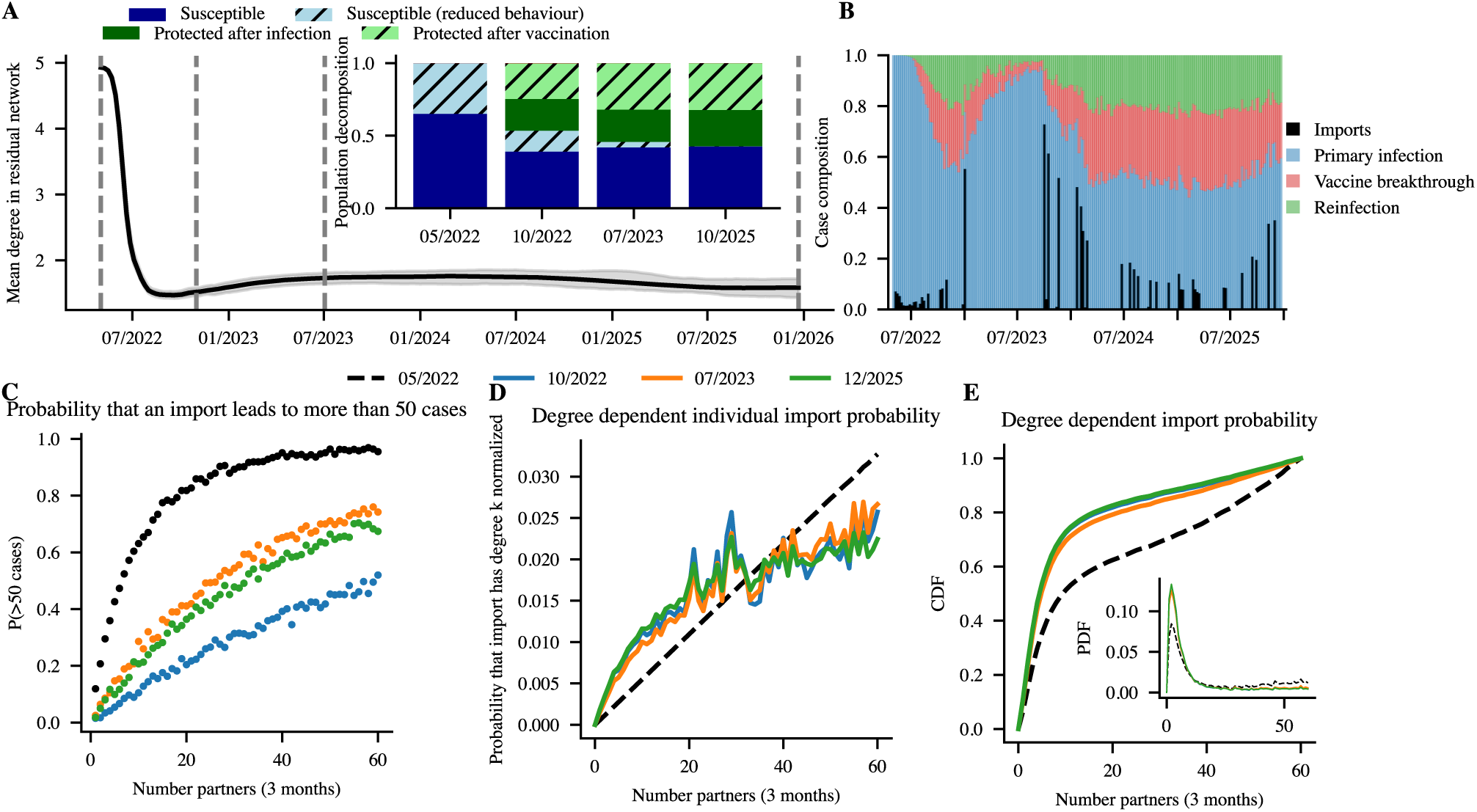
Population immunity gaps. Analogue of Fig. 3 (main manuscript) for model M2. **(A)** Mean contact degree of susceptibles over time. Grey shading indicates the 95% CI. Vertical lines mark the start of the simulation (05/2022), the end of the calibration period (10/2022), immediately before mpox re-emergence in Berlin (07/2023), and the end of the simulation (12/2025). The inset shows the population composition by protection and behavioural status at these four time points. **(B)** Weekly case composition by infection type (primary infection, vaccine-breakthrough infection, and reinfection), with imported cases shown separately (black bars). **(C)** Probability that a single infection (e.g. an imported case) of an individual with *k* contacts generates an outbreak with more than 50 secondary cases. For each degree and time point, 1,000 simulations were performed. **(D)** Probability that an infection (e.g. importation) occurs in an individual with *k* contacts, normalised by the size of the corresponding degree class, at the four time points. **(E)** Probability that an imported case has degree *k*. The inset shows the probability density function, and the main panel shows the cumulative distribution function.

**Supplementary Figure S5:**
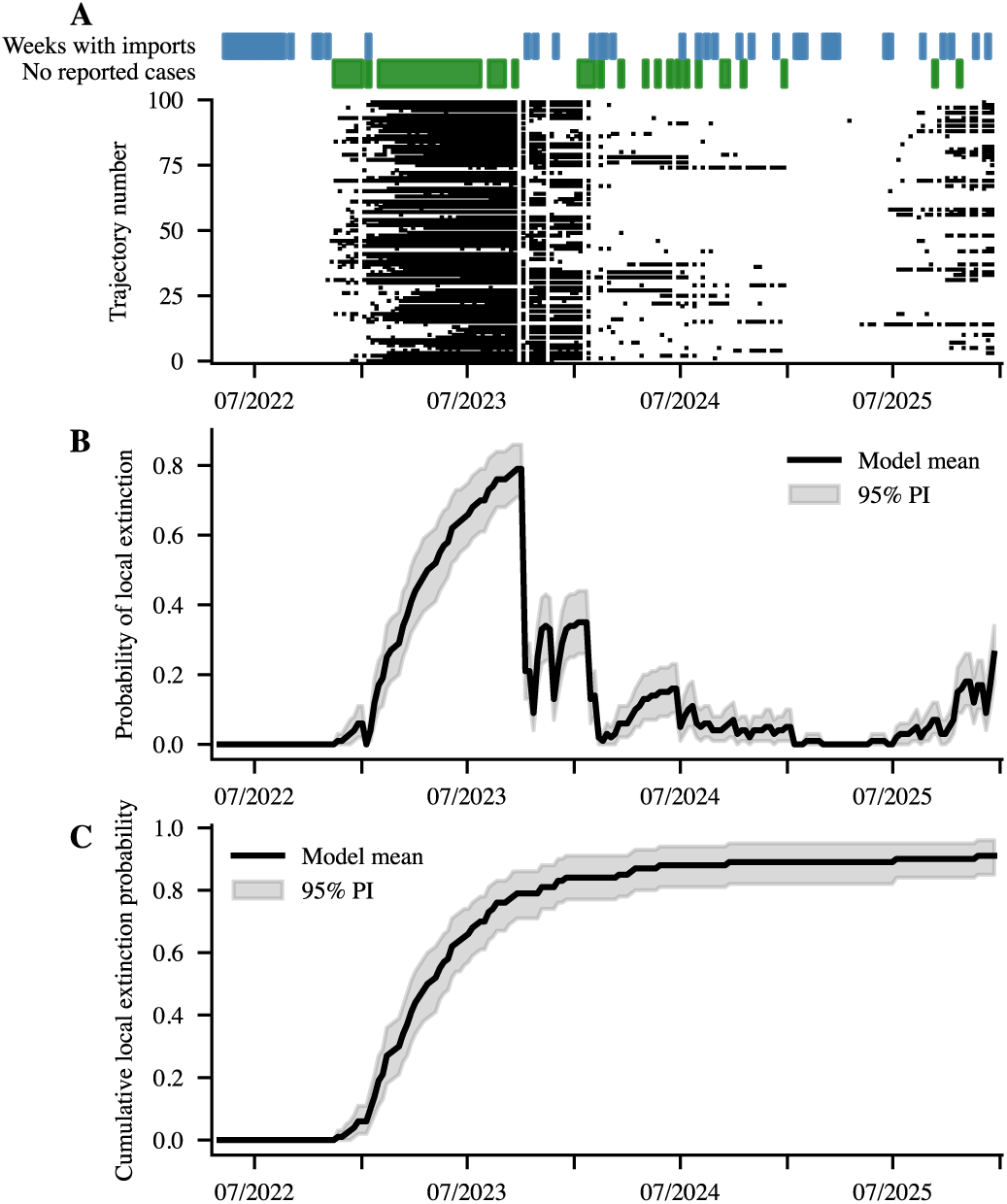
Temporal case composition and extinction probability Analogue of Fig. 4 (main manuscript) for model M2. **(A)** Green bars indicate weekly periods with no reported cases in Berlin/Germany and the blue bars indicate weeks were cases were imported. Base on 100 horizontally plotted stochastic simulations we depicted black brigs if no endogenous infections occurred within a week. **(B)** Weekly probability of no active infections (i.e., extinction) for each model (i.e. vertically counting the frequency of ‘black brigs’ from panel A). Solid lines show the mean across bootstrap replicates and shaded bands the 95% bootstrap confidence interval (CI). **(C)** Cumulative probability of observing at least one week with no active infections by a given week (i.e. horizontally counting if a ‘black brigs’ appeared in panel A).

**Supplementary Figure S6:**
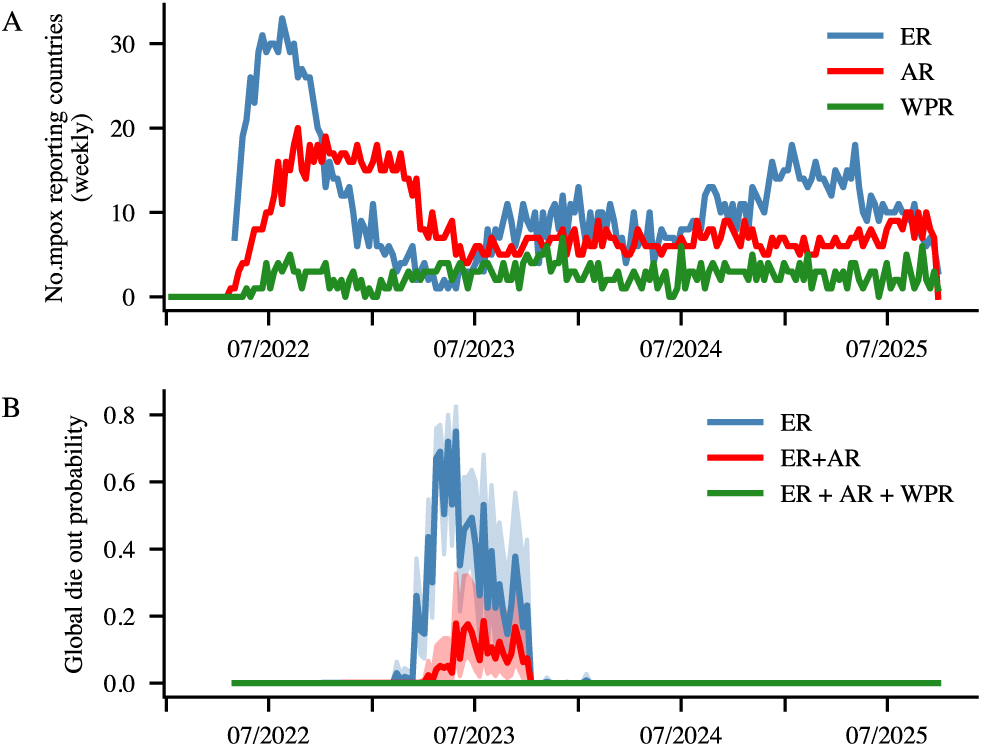
Asynchronous global outbreaks reduce mpox extinction probability. Analogue of Fig. 5 (main manuscript) for model M2. Estimated extinction probability for mpox, calculated for combinations of regions: European region alone (blue); European and American regions (red); and European, American, and Western Pacific regions (green). Estimates are based on Berlin-derived weekly extinction probabilities applied to regional surveillance data.

**Supplementary Figure S7:**
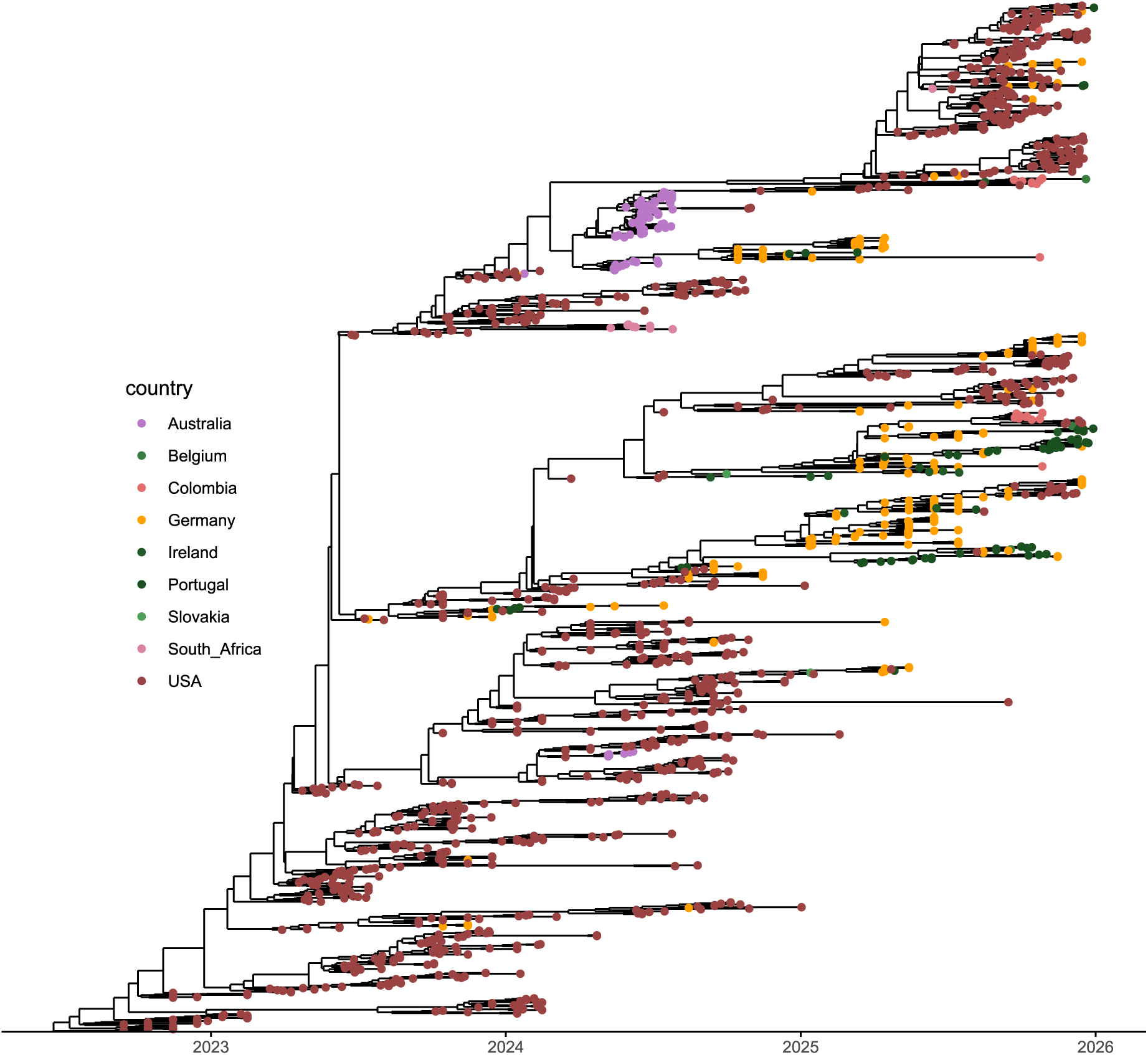
BEAST2 timetree of sequences belonging to clades B.1.20, which includes active clusters F.2 and F.4 with country of origin indicated.

**Supplementary Figure S8:**
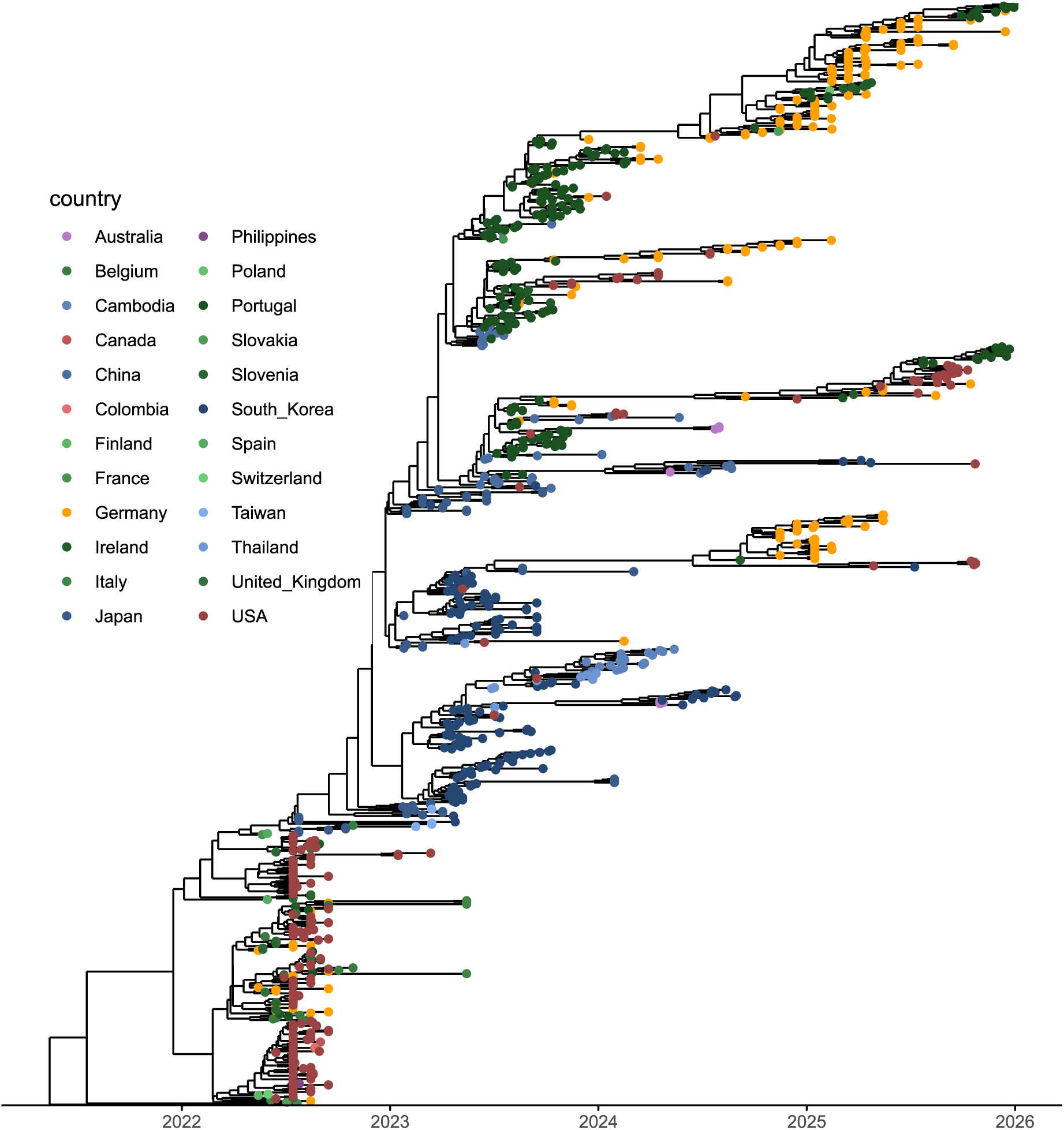
BEAST2 timetree of sequences belonging to clades B.1.3, which includes active clusters C.1.2, E.1.1, E.2 and E.3.1 with country of origin indicated.

**Supplementary Figure S9:**
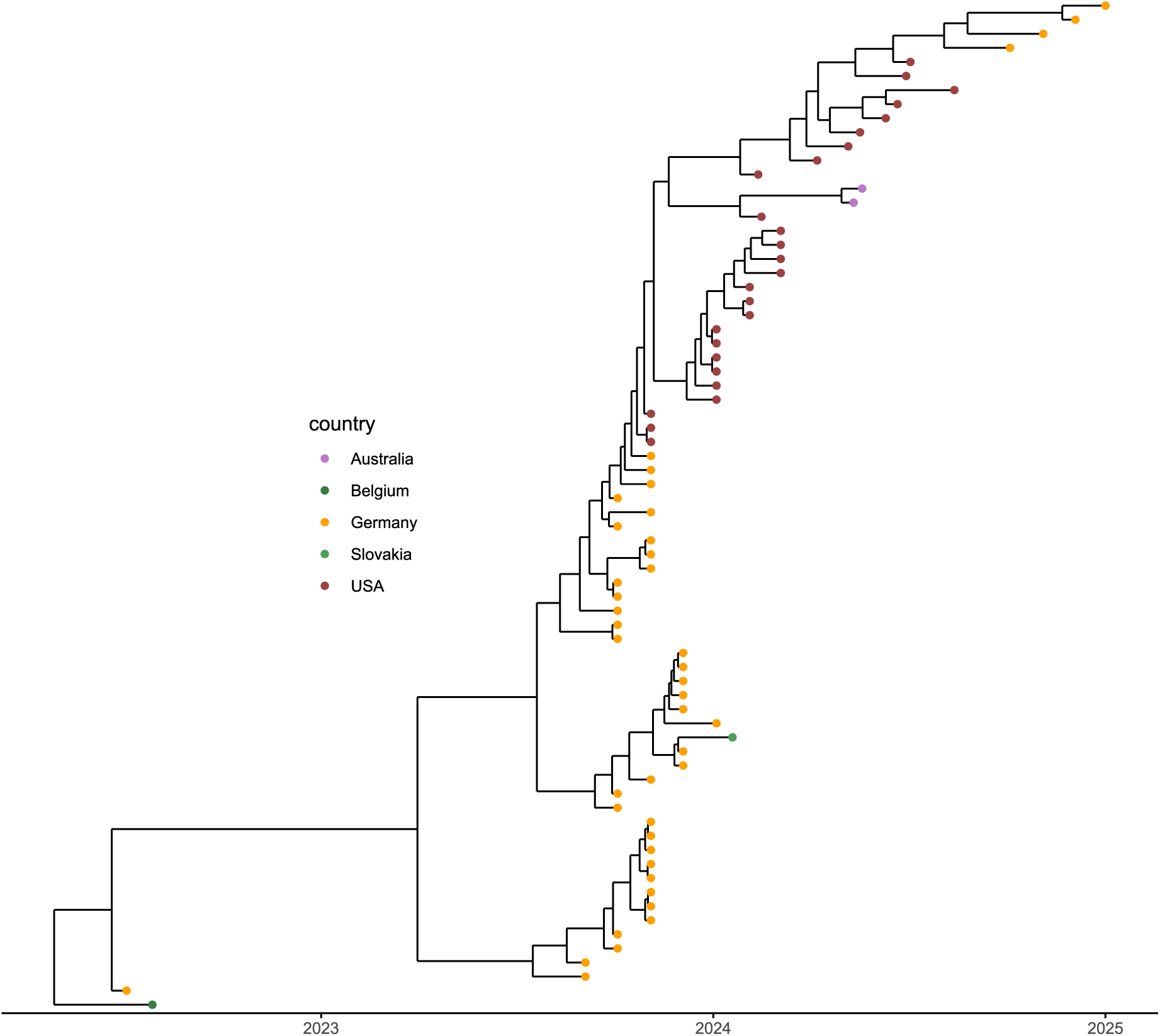
BEAST2 timetree of sequences belonging to clades B.1.22, which includes subclade J.1 with country of origin indicated.

**Supplementary Figure S10:**
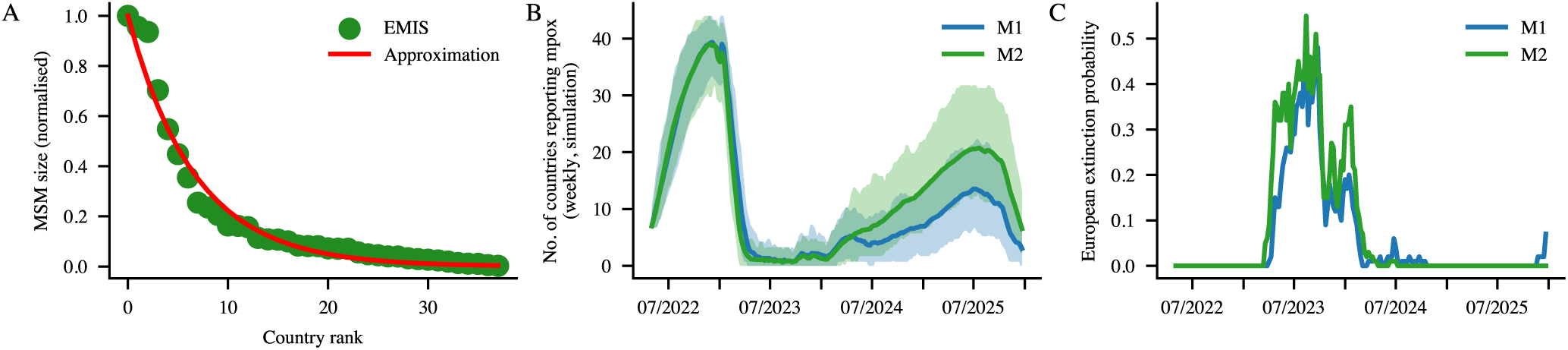
Metapopulation Simulation for the European Region. **(A)** Distribution of MSM population sizes in Europe, as estimated by EMIS (indicated by green dots), using an exponential distribution (red line) with a parameter of 0.15. **(B)** Modelderived number of European countries reporting mpox cases per week. **(C)** Model-derived extinction probability of mpox in Europe.

**Supplementary Table S1:**
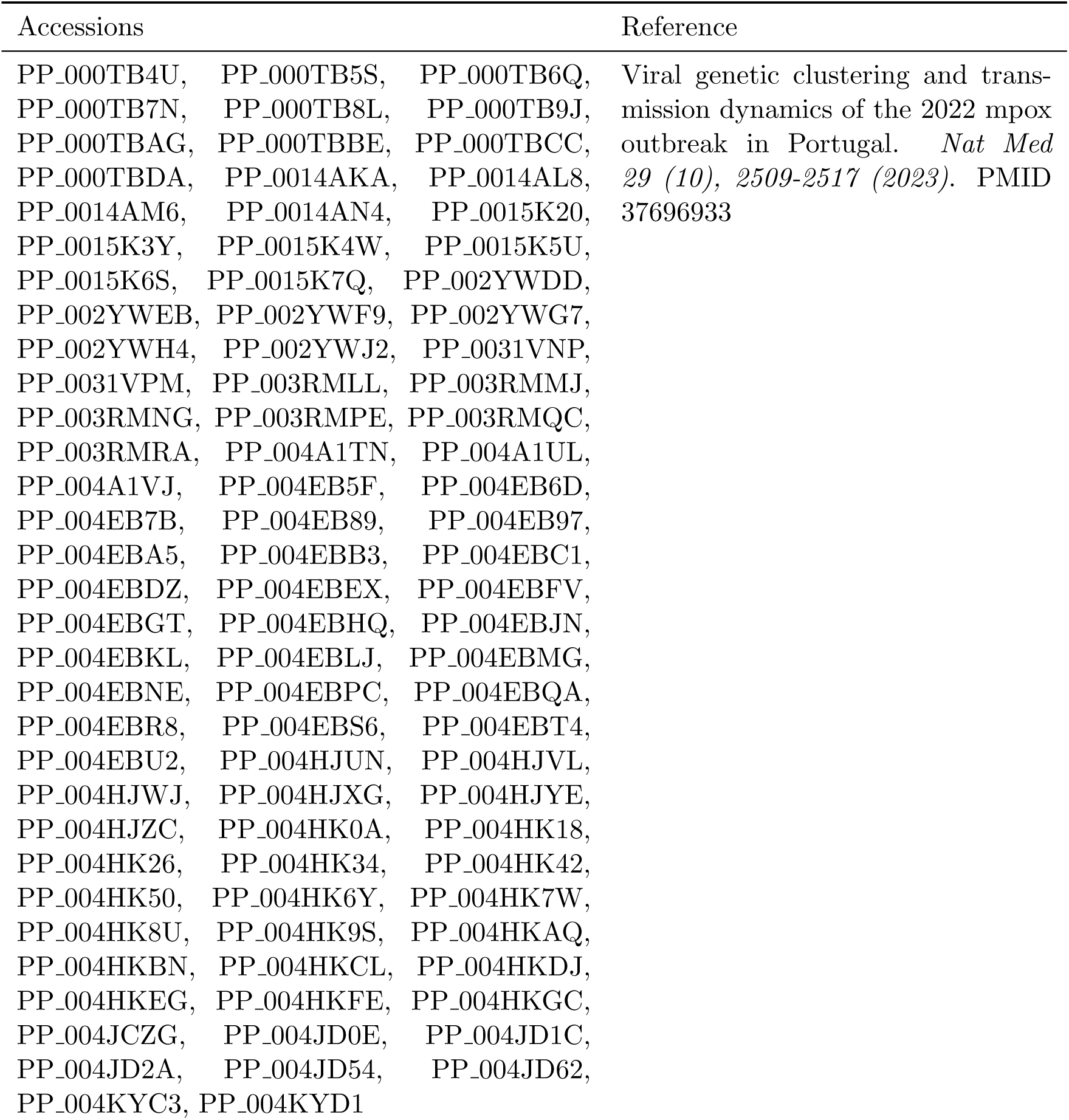
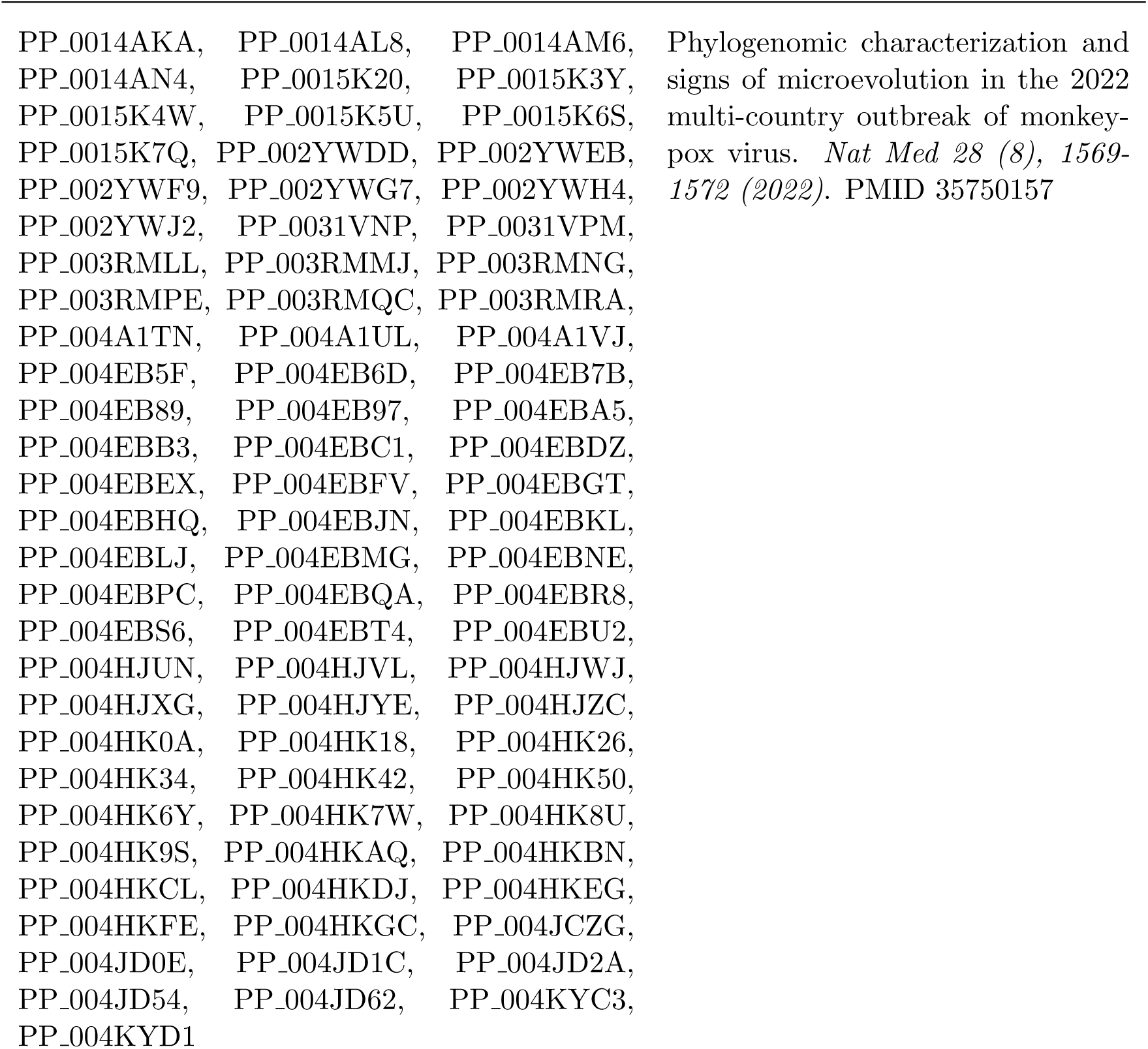
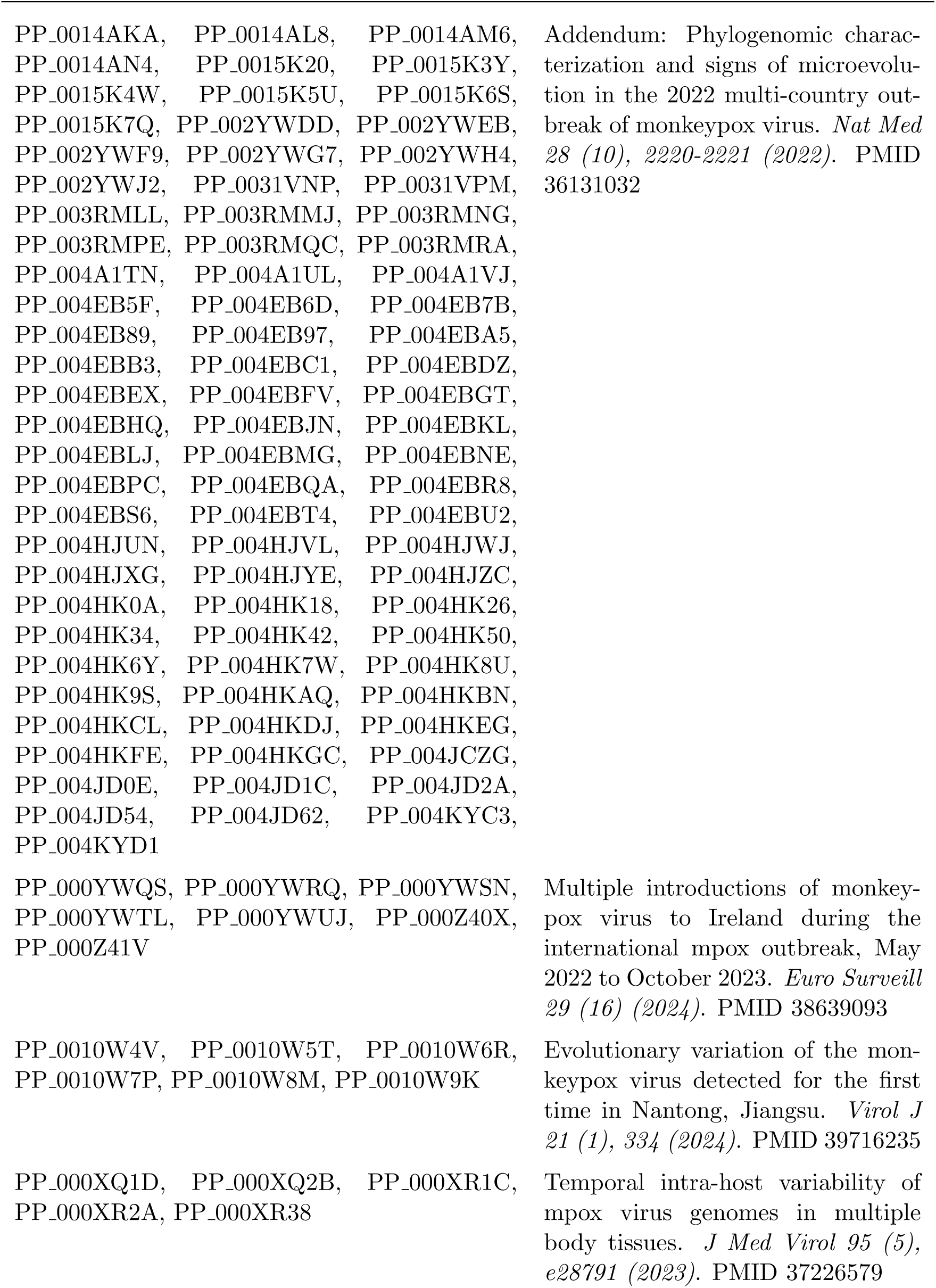
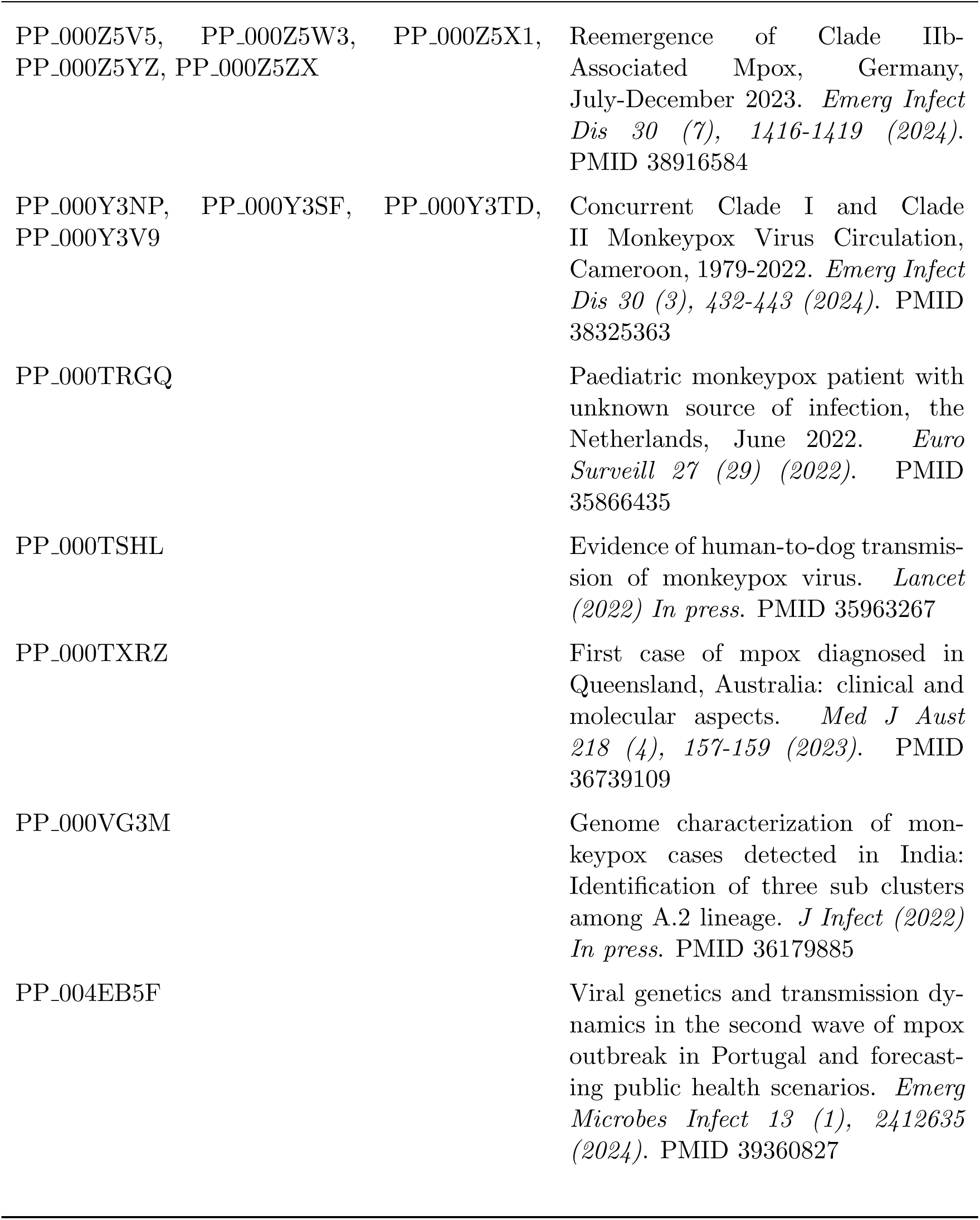
Sequences used in this study for which a publication is available.

**Supplementary Table S2:**
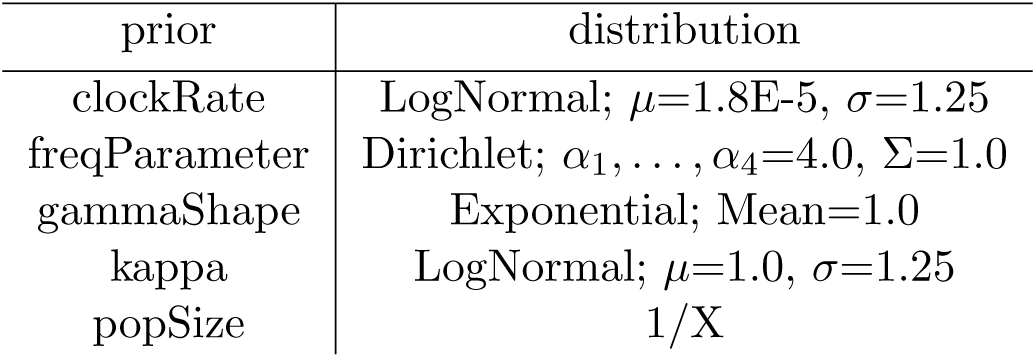
Priors used in BEAST2.

Supplementary Table ST2: Priors used in BEAST2
| prior | distribution |
| --- | --- |
| clockRate | LogNormal; $\mu=1.8\text{E-}5$, $\sigma=1.25$ |
| freqParameter | Dirichlet; $\alpha_1, \dots, \alpha_4=4.0$, $\Sigma=1.0$ |
| gammaShape | Exponential; Mean=1.0 |
| kappa | LogNormal; $\mu=1.0$, $\sigma=1.25$ |
| popSize | $1/X$ |

## References

[1] Dimie Ogoina, Jake Dunning, Inger Damon, Placide Mbala, and Krutika Kuppalli. Mpox emergence, epidemiology, biology, clinical features and control. Nature Reviews Microbiology, pages 1–15, 2026.

[2] Nicola L Bragazzi, Jude D Kong, Naim Mahroum, Christina Tsigalou, Rola Khamisy-Farah, Manlio Converti, and Jianhong Wu. Epidemiological trends and clinical features of the ongoing monkeypox epidemic: a preliminary pooled data analysis and literature review. Journal of medical virology, 95(1):e27931, 2023.

[3] Nicola Luigi Bragazzi, Jude Dzevela Kong, and Jianhong Wu. Is monkeypox a new, emerging sexually transmitted disease? a rapid review of the literature. Journal of Medical Virology, 95(1):e28145, 2023.

[4] Pengfei Li, Jiajing Li, Ibrahim Ayada, Amine Avan, Qinyue Zheng, Maikel P Peppelenbosch, Annemarie C de Vries, and Qiuwei Pan. Clinical features, antiviral treatment, and patient outcomes: a systematic review and comparative analysis of the previous and the 2022 mpox outbreaks. The Journal of Infectious Diseases, 228(4):391–401, 2023.

[5] Ulrik Hvid, Lone Simonsen, Morten Frisch, and Kim Sneppen. Relationship dynamics and behavioral adaptations in the control of the 2022 mpox epidemic. Proceedings of the National Academy of Sciences, 122(37):e2502861122, 2025.

[6] Giorgio Guzzetta, Alessia Mammone, Federica Ferraro, Anna Caraglia, Alessia Rapiti, Valentina Marziano, Piero Poletti, Danilo Cereda, Francesco Vairo, Giovanna Mattei, et al. Early estimates of monkeypox incubation period, generation time, and reproduction number, italy, may–june 2022. Emerging infectious diseases, 28(10):2078, 2022.

[7] Maria Xiridou, Fuminari Miura, Philippe Adam, Eline Op de Coul, John de Wit, and Jacco Wallinga. The fading of the mpox outbreak among men who have sex with men: A mathematical modelling study. The Journal of Infectious Diseases, 230(1):e121–e130, 07 2024.

[8] Patrick A Clay, Jason M Asher, Neal Carnes, Casey E Copen, Kevin P Delaney, Daniel C Payne, Emily D Pollock, Jonathan Mermin, Yoshinori Nakazawa, William Still, Anil T Mangla, and Ian H Spicknall. Modelling the impact of vaccination and sexual behaviour adaptations on mpox cases in the usa during the 2022 outbreak. Sexually Transmitted Infections, 100(2):70–76, 2024.

[9] Elise De Vos, Liesbeth Van Gestel, Isabel Brosius, Chris Kenyon, Bea Vuylsteke, Irith De Baetselier, Joachim Marien, Eugene Bangwen, Simon Couvreur, Amaryl Lecompte, et al. Potential determinants of the decline in mpox cases in belgium: A behavioral, epidemiological and seroprevalence study. International Journal of Infectious Diseases, 146:107132, 2024.

[10] Nils Gubela, Hee-yeong Kim, Nikolay Lunchenkov, Daniel Stern, Janine Michel, Andreas Nitsche, Axel J Schmidt, Ulrich Marcus, and Max von Kleist. Behavior change and infection induced immunity led to the decline of the 2022 mpox outbreak in berlin. Communications medicine, 6(1):81, 2026.

[11] Davide Maniscalco, Olivier Robineau, Pierre-Yves Boëlle, Mattia Mazzoli, Anne-Sophie Barret, Emilie Chazelle, Alexandra Mailles, Harold Noël, Arnaud Tarantola, Annie Velter, et al. Role of behaviour change in controlling the 2022 paris mpox outbreak. Nature Health, pages 1–12, 2026.

[12] Aisling M Vaughan, Mohammed Afzal, Priyanka Nannapaneni, Mathias Leroy, Xanthi Andrianou, Jeffrey Pires, Silvia Funke, Celine Roman, Juliana Reyes-Uruena, Stephan Aberle, Aristos Aristodimou, Gudrun Aspelund, Kirsty F Bennet, Antra Bormane, Anna Caraglia, Hannah Charles, Emilie Chazelle, Iva Christova, Orna Cohen, Costas Constantinou, Simon Couvreur, Asuncion Diaz, Kateřina Fabiánová, Federica Ferraro, Marte Petrikke Grenersen, Eva Grilc, Tuula Hannila-Handelberg, Anne Kathrine Hvass, Derval Igoe, Klaus Jansen, Denisa Janţă, Styliani Kaoustou, Anders Koch, Mirjana Lana Kosanovic Licina, Stefka Krumova, Anton Labutin, Raskit Lachmann, Amaryl Lecompte, Rémi Lefrançois, Viktorija Leitena, Kirsi Liitsola, Ivan Mlinarić, Zohar Mor, Martha Neary, Alina Novacek, Magnus Wenstøp Øgle, Hana Orlıková, Kalliopi Papadima, Moa Rehn, Malgorzata Sadkowska-Todys, Anca Sîrbu, Klara Sondén, Berta Suárez, Marianna Thordardottir, Paula Vasconcelos, Joao Vieira Martins, Karolina Zakrzewska, Marc-Alain Widdowson, and Céline M Gossner. Continued circulation of mpox: an epidemiological and phylogenetic assessment, european region, 2023 to 2024. Eurosurveillance, 29(27), 2024.

[13] Clara Suñer, Maria Ubals, Eloy José Tarín-Vicente, Adrià Mendoza, Andrea Alemany, Águeda Hernández-Rodrıguez, Cristina Casañ, Vicente Descalzo, Dan Ouchi, Aurélien Marc, et al. Viral dynamics in patients with monkeypox infection: a prospective cohort study in spain. The Lancet Infectious Diseases, 23(4):445–453, 2023.

[14] Silvia Meschi, Francesca Colavita, Fabrizio Carletti, Valentina Mazzotta, Giulia Matusali, Eliana Specchiarello, Tommaso Ascoli Bartoli, Annalisa Mondi, Claudia Minosse, Maria Letizia Giancola, Carmela Pinnetti, Maria Beatrice Valli, Daniele Lapa, Klizia Mizzoni, David J Sullivan, Jiangda Ou, Daniele Focosi, Enrico Girardi, Emanuele Nicastri, Andrea Antinori, and Fabrizio Maggi. MPXV DNA kinetics in bloodstream and other body fluids samples. Scientific Reports, 14(1):13487, June 2024.

[15] Miguel I Paredes, Citina Liang, Sze-chuan Suen, Ian W Holloway, Jacob M Garrigues, Nicole M Green, Trevor Bedford, Nicola F Müller, and Joseph Osmundson. Viral introductions and return to baseline sexual behaviors maintain low-level mpox incidence in los angeles. Nature Communications, 2026.

[16] Akira Endo, Hiroaki Murayama, Sam Abbott, Ruwan Ratnayake, Carl AB Pearson, W John Edmunds, Elizabeth Fearon, and Sebastian Funk. Heavy-tailed sexual contact networks and monkeypox epidemiology in the global outbreak, 2022. Science, 378(6615):90–94, 2022.

[17] Joseph A Lewnard, Miguel I Paredes, Matan Yechezkel, Gregg S Davis, Vennis Hong, Jessica Skela, Utsav Pandey, Noah T Parker, Lauren C Granskog, Magdalena E Pomichowski, et al. Extensive cryptic circulation sustains mpox among men who have sex with men. Nature Communications, 17(1):4198, 2026.

[18] Luca M Zaeck, Mart M Lamers, Babs E Verstrepen, Theo M Bestebroer, Martin E Van Royen, Hannelore Götz, Marc C Shamier, Leanne PM Van Leeuwen, Katharina S Schmitz, Kimberley Alblas, et al. Low levels of monkeypox virus-neutralizing antibodies after mva-bn vaccination in healthy individuals. Nature medicine, 29(1):270–278, 2023.

[19] Matthew T Berry, Shanchita R Khan, Timothy E Schlub, Adriana Notaras, Mohana Kunasekaran, Andrew E Grulich, C Raina MacIntyre, Miles P Davenport, and David S Khoury. Predicting vaccine effectiveness for mpox. Nature Communications, 15(1):3856, 2024.

[20] Tingting Li, Zhijin Li, Yu Xia, Jiang Long, and Li Qi. Mpox reinfection: A rapid systematic review of case reports. Infectious Medicine, 3(1):100096, 2024.

[21] Aniruddha Hazra, Jason Zucker, Elizabeth Bell, John Flores, Leanna Gordon, Oriol Mitjà, Clara Suñer, Adrien Lemaignen, Simon Jamard, Silvia Nozza, et al. Mpox in people with past infection or a complete vaccination course: a global case series. The Lancet Infectious Diseases, 24(1):57–64, 2024.

[22] Robert Koch-Institut. Survstat@rki 2.0. https://survstat.rki.de. Accessed: 2025-01-25.

[23] World Health Organization (WHO). 2022-26 Mpox Outbreak: Global Trends. https://worldhealthorg.shinyapps.io/mpx_global/, 2026. Accessed: 2026-05-22.

[24] Patrick E Obermeier, Clarissa F Plinke, Annika Brinkmann, Raskit Lachmann, Julia Melchert, Victor M Corman, Andreas Nitsche, Ulrich Marcus, Axel J Schmidt, Klaus Jansen, et al. Reemergence of clade iib–associated mpox, germany, july–december 2023. Emerging Infectious Diseases, 30(7):1416, 2024.

[25] Nils Gubela and Max von Kleist. Efficient and accurate simulation of infectious diseases on adaptive networks. PLOS Complex Systems, 2(6):e0000049, 2025.

[26] Ulrich Marcus and Susanne Schink. Empirische ergebnisse und handlungsempfehlungen zur hiv-/sti-prävention und gesundheitsförderung bei schwulen und bisexuellen männern in deutschland. EMIS: The European MSM-Internet-Survey 2017, 2022.

[27] Faisal S Minhaj, Vijay Singh, Stephanie E Cohen, Michael B Townsend, Hyman Scott, John Szumowski, C Bradley Hare, Pallavi Upadhyay, Jairus Reddy, Barbara Alexander, et al. Prevalence of undiagnosed monkeypox virus infections during global mpox outbreak, united states, june–september 2022. Emerging infectious diseases, 29(11):2307, 2023.

[28] Stefano Musumeci, Jérôme Laflamme, Laurent Kaiser, Olivier Segeral, and Alexandra Calmy. Characteristics of possible mpox reinfection cases: literature review. Journal of Travel Medicine, 30(7):taad136, 2023.

[29] Anaís Corma-Gómez, Alfonso Cabello, Eva Orviz, Miguel Morante-Ruiz, Oskar Ayerdi, Aws Al-Hayani, Ana Munõz-Gómez, Ignacio De Los Santos, Cristina Gómez-Ayerbe, David Rodrigo, et al. Long or complicated mpox in patients with uncontrolled hiv infection. Journal of Medical Virology, 96(3):e29511, 2024.

[30] Giorgio Guzzetta, Valentina Marziano, Alessia Mammone, Andrea Siddu, Federica Ferraro, Anna Caraglia, Francesco Maraglino, Giovanni Rezza, Alessandro Vespignani, Ira Longini, et al. The decline of the 2022 italian mpox epidemic: Role of behavior changes and control strategies. Nature Communications, 15(1):2283, 2024.

[31] Fanyu Xiu, Carla Doyle, Jorge Luis Flores Anato, Jesse Knight, Linwei Wang, Joseph Cox, Daniel Grace, Trevor A Hart, Terri Zhang, Shayna Skakoon-Sparling, et al. Impact of interventions on mpox transmission during the 2022 outbreak in canada: a mathematical modeling study of three different cities. International Journal of Infectious Diseases, page 107792, 2025.

[32] Matthew J Ferrari, Shweta Bansal, Lauren A Meyers, and Ottar N Bjørnstad. Network frailty and the geometry of herd immunity. Proceedings of the Royal Society B: Biological Sciences, 273(1602):2743–2748, 08 2006.

[33] European Centre for Disease Prevention and Control & WHO Regional Office for Europe. Joint ECDC-WHO Regional Office for Europe Mpox Surveillance Bulletin. https://monkeypoxreport.ecdc.europa.eu, 2026. Accessed: 2025-12-01.

[34] Akira Endo, Sung-Mok Jung, and Fuminari Miura. Mpox emergence in japan: ongoing risk of establishment in asia. The Lancet, 401(10392):1923–1924, 2023.

[35] Mingyeol Shim, Soo Hyeon Cho, Seung Eun Lee, and Taeyoung Kim. Epidemiological characteristics and risk factors of suspected and confirmed mpox cases during the 2022– 2023 epidemic in the capital region, korea. Epidemiology and Health, 46:e2024092, 2024.

[36] Ting Xu and Leiliang Zhang. Rising prevalence of mpox in china, japan, and republic of korea. Journal of Infection, 87(4):e73–e74, 2023.

[37] Elena Dalla Vecchia. Pathoplexus: towards fair and transparent sequence sharing. The Lancet. Microbe, 5:100995, December 2024.

[38] James Hadfield, Colin Megill, Sidney M. Bell, John Huddleston, Barney Potter, Charlton Callender, Pavel Sagulenko, Trevor Bedford, and Richard A. Neher. Nextstrain: realtime tracking of pathogen evolution. Bioinformatics (Oxford, England), 34:4121–4123, December 2018.

[39] Hiroaki Murayama, Carl AB Pearson, Sam Abbott, Fuminari Miura, Sung-mok Jung, Elizabeth Fearon, Sebastian Funk, and Akira Endo. Accumulation of immunity in heavy-tailed sexual contact networks shapes mpox outbreak sizes. The Journal of infectious diseases, 229(1):59–63, 2024.

[40] Romualdo Pastor-Satorras, Claudio Castellano, Piet Van Mieghem, and Alessandro Vespignani. Epidemic processes in complex networks. Rev. Mod. Phys., 87:925–979, Aug 2015.

[41] Bo Peng, Ziquan Lyu, Wenxiao Gong, Xiaomin Zhang, Shiting Chen, Wenjuan Ma, Xiaolu Shi, Jia Wan, Jing Qu, and Xiaoping Dong. Emergence of lineage e.4 and structural plasticity of a28l protein in mpox virus: Characterization of lcr7 length polymorphisms across lineages shenzhen city, guangdong province, china, 2023-2025. China CDC weekly, 8:847–851, July 2026.

[42] Chuan Shi, Xiaochen Zheng, Lei Lei, Jinhui Xiao, Guangqing Yu, Yingdong Li, Zhifeng Ma, Minjie Li, Yanling Zeng, Ziquan Lv, Yixiong Chen, Wei Tan, and Qianru Wang. Phylogenetic and molecular evolutionary insights into monkeypox virus circulation in shenzhen, china, 2023-2024. Viruses, 17, September 2025.

[43] Chengyunxiao Li, Dali Xu, Qing Duan, Hao Wang, Yan Li, Shujun Ding, Ti Liu, Renpeng Li, Zengqiang Kou, and Chunhong Yin. Evolutionary epidemiology of the monkeypox virus in shandong province during the post-global outbreak era. Frontiers in microbiology, 16:1677051, 2025.

[44] Rita Cordeiro, Constantino P Caetano, Daniel Sobral, Rita Ferreira, Luís Coelho, Ana Pelerito, Isabel Lopes de Carvalho, Sónia Namorado, Dinis B Loyens, Ricardo Mexia, et al. Viral genetics and transmission dynamics in the second wave of mpox outbreak in portugal and forecasting public health scenarios. Emerging Microbes & Infections, 13(1):2412635, 2024.

[45] Shengjie Zhang, Fuxiang Wang, Yun Peng, Xiaohua Gong, Guohao Fan, Yuanlong Lin, Liuqing Yang, Liang Shen, Shiyu Niu, Jiexiang Liu, et al. Evolutionary trajectory and characteristics of mpox virus in 2023 based on a large-scale genomic surveillance in shenzhen, china. Nature Communications, 15(1):7452, 2024.

[46] Ranran Cao, Chengxia Liu, Ying Shi, Can Luo, Xin Chen, Li Liu, Chaohua Xu, Ming Pan, Changcheng Wu, Li Zhang, and Wenjie Tan. Genomic surveillance and phylogenetic analysis of monkeypox virus sampled from clinical monkeypox cases and sewage -sichuan province, china, 2023. China CDC weekly, 7:1182–1191, September 2025.

[47] Dennis D Taub, William B Ershler, Mark Janowski, Andrew Artz, Michael L Key, Julie McKelvey, Denis Muller, Bernard Moss, Luigi Ferrucci, Patricia L Duffey, et al. Immunity from smallpox vaccine persists for decades: a longitudinal study. The American journal of medicine, 121(12):1058–1064, 2008.

[48] N. Alexia Raharinirina, Nils Gubela, Daniela Börnigen, Maureen Smith, Djin-Ye Oh, Matthias Budt, Christin Mache, Claudia Schillings, Stephan Fuchs, Ralf Dürrwald, Thorsten Wolff, Martin Hölzer, Sofia Paraskevopoulou, and Max von Kleist. Sars-cov-2 evolution on a dynamic immune landscape. Nature, 639:196–204, 2025.

[49] Matthijs Meijers, Denis Ruchnewitz, Jan Eberhardt, Marta Luksza, and Michael Lässig. Population immunity predicts evolutionary trajectories of sars-cov-2. Cell, 186:5151–5164.e13, November 2023.

[50] Marta Luksza and Michael Lassig. A predictive fitness model for influenza. Nature, 507:57–61, March 2014.

[51] Martha I. Nelson, Lone Simonsen, Cecile Viboud, Mark A. Miller, and Edward C. Holmes. Phylogenetic analysis reveals the global migration of seasonal influenza a viruses. PLoS pathogens, 3:1220–1228, September 2007.

[52] Ulrich Marcus, Janine Michel, Nikolay Lunchenkov, Denis Beslic, Fridolin Treindl, Rebecca Surtees, Christoph Weber, Axel Baumgarten, Andreas Nitsche, and Daniel Stern. A seroprevalence study indicates a high proportion of clinically undiagnosed mpxv infections in men who have sex with men in berlin, germany. BMC infectious diseases, 24(1):1153, 2024.

[53] Sarah Adamo, Yu Gao, Takuya Sekine, Akhirunnesa Mily, Jinghua Wu, Elisabet Storgärd, Victor Westergren, Finn Filen, Carl-Johan Treutiger, Johan K Sandberg, et al. Memory profiles distinguish cross-reactive and virus-specific t cell immunity to mpox. Cell host & microbe, 31(6):928–936, 2023.

[54] Martina Rueca, Fabio Giovanni Tucci, Valentina Mazzotta, Giulia Gramigna, Cesare Ernesto Maria Gruber, Lavinia Fabeni, Emanuela Giombini, Giulia Matusali, Carmela Pinnetti, Andrea Mariano, et al. Temporal intra-host variability of mpox virus genomes in multiple body tissues. Journal of Medical Virology, 95(5):e28791, 2023.

[55] Samuel Schildhauer, Kayla Saadeh, Robert E Snyder, Eric C Tang, Eric Chapman, Deanna A Sykes, Philip Peters, Kathleen Jacobson, Jessica Watson, and Kelly A Johnson. Prolonged monkeypox virus infections, california, usa, may 2022–august 2024. Emerging Infectious Diseases, 31(10):1935, 2025.

[56] Matthias an der Heiden, Ulrich Marcus, Christian Kollan, Daniel Schmidt, Barbara Gunsenheimer-Bartmeyer, and Viviane Bremer. Schätzung der zahl der hivneuinfektionen und dergesamtzahl von menschen mit hiv in deutschland,stand ende 2020. Epid Bull, 47:3–17, 2021.

[57] Kevin L Karem, Mary Reynolds, Christine Hughes, Zach Braden, Pragati Nigam, Shane Crotty, John Glidewell, Rafi Ahmed, Rama Amara, and Inger K Damon. Monkeypoxinduced immunity and failure of childhood smallpox vaccination to provide complete protection. Clinical and Vaccine Immunology, 14(10):1318–1327, 2007.

[58] Michael Thy, Nathan Peiffer-Smadja, Morgane Mailhe, Laura Kramer, Valentine M. Ferré, Nadhira Houhou, Hassan Tarhini, Chloé Bertin, Anne-Lise Beaumont, Mathilde Garé, Diane Le Pluart, Ségolène Perrineau, Mayda Rahi, Laurène Deconinck, Bao Phung, Bastien Mollo, Marie Cortier, Mélanie Cresta, Clémentine De La Porte Des Vaux, Véronique Joly, Sylvie Lariven, Christophe Rioux, Cécile Somarriba, FrancoisXavier Lescure, Charlotte Charpentier, Yazdan Yazdanpanah, and Jade Ghosn. Breakthrough infections after postexposure vaccination against mpox. New England Journal of Medicine, 387(26):2477–2479, 2022.

[59] Hao Liu, Wenjing Wang, Yang Zhang, Fuchun Wang, Junyi Duan, Tao Huang, Xiaojie Huang, and Tong Zhang. Global perspectives on smallpox vaccine against monkeypox: a comprehensive meta-analysis and systematic review of effectiveness, protection, safety and cross-immunogenicity. Emerging Microbes & Infections, 13(1):2387442, 2024.

[60] Alexander Bartel, Klaus Jansen, Ronja Boberg, Julia Bitzegeio, Annika Brinkmann, Livia Schrick, Raskit Lachmann, Daniel Sagebiel, Andreas Nitsche, Claudia Ruscher, et al. Rapid spread of mpxv clade ib with high genetic relatedness among men who have sex with men, berlin, germany, week 50 2025 up to week 10 2026. Eurosurveillance, 31(12):2600235, 2026.

[61] Klaus Jansen, Judith Koch, Gyde Steffen, and Raskit Lachmann. Aktuelle epidemiologische situation von mpox in deutschland. Epid Bull, pages 13–15, 2026.

[62] Fuminari Miura, Ka Yin Leung, Maria Xiridou, Marten van Antwerpen, Nicola Low, Niel Hens, Emmanuel Hasivirwe Vakaniaki, and Jacco Wallinga. Dynamic shift in the dominant transmission route of clade ib monkeypox virus across networks with sexual and nonsexual contacts. Science advances, 12:eaec1931, April 2026.

[63] Majid M. Alshamrani, Aiman El-Saed, Sarah Al-Fayez, Kholod AlAmeer, Mohammed Al Zunitan, Mohammed Abalkhail, Fatmah Othman, Fayssal Farahat, Syed Nazeer, Wafaa Al Nasser, Maher Alharbi, Tom Fletcher, Tochi Okwor, Hibak Mahamed, Hannah Hamilton Hurwitz, Victoria Willet, and April Baller. Routes of transmission of mpox by virus clade and geographic distribution: A systematic review. Journal of infection and public health, 18:102985, December 2025.

[64] Jameson Crandell, Raianna F. Fantin, Leonardo Pereira de Araújo, Lauren Lawres, Lauren Pischel, Valter S. Monteiro, Luciana Conde, Angelica C. Kottkamp, Marie I. Samanovic, Rita Cordeiro, Diana Póvoas, Mariana Melo, Terezinha M. Castineiras, Andre M. Vale, Inci Yildirim, Mark J. Mulligan, Leonardo Augusto de Almeida, Saad B. Omer, Camila H. Coelho, and Carolina Lucas. Neutralising antibody responses to mpxv clades ia, ib, and iib after infection or vaccination: a multicountry observational study. The Lancet. Infectious diseases, June 2026.

[65] Jing Liu, Xun Wang, Yiting Zhang, Changyi Liu, Meng Zhang, Chen Li, Peiling Liu, Shanshan Li, Kaifeng Wei, Yiming Cai, Hongjie Yu, Zhiliang Hu, Pengfei Wang, and Yanliang Zhang. Immunogenicity of monkeypox virus surface proteins and cross-reactive antibody responses in vaccinated and infected individuals: implications for vaccine and therapeutic development. Infectious diseases of poverty, 14:12, February 2025.

[66] Jan Stratil, Alexandra Hofmann, Viviane Bremer, Anette Siedler, Klaus Jansen, and Uwe Koppe. Aufbau, struktur und ergebnisse eines freiwilligen mpox-impfmonitorings in deutschland. Epid Bull, 43:3–12, 2023.

[67] Romualdo Pastor-Satorras, Claudio Castellano, Piet Van Mieghem, and Alessandro Vespignani. Epidemic processes in complex networks. Rev. Mod. Phys., 87:925–979, Aug 2015.

[68] Joanne Byrne, Alejandro Garcia-Leon, Aisling Murphy, Gurvin Saini, Ishan Banik, Alan Landay, Liem Binh Luong Nguyen, Stefano Savinelli, Cathal O’Broin, Mary Horgan, Christine Kelly, Carlos Mejia-Chew, Corinna Sadlier, Eoghan de Barra, Jane A O’Halloran, Virginie Gautier, Patrick W G Mallon, and on behalf of the All Ireland Infectious Diseases Cohort Study Feeney, Eoin R. Antibody responses are sustained 2 years post-mpox infection but not following modified vaccinia ankara–bavarian nordic vaccination. Open Forum Infectious Diseases, 12(9):ofaf536, 09 2025.

[69] Marta Bertran, Nick Andrews, Chloe Davison, Bennet Dugbazah, Jacob Boateng, Rachel Lunt, Joanne Hardstaff, Melanie Green, Paula Blomquist, Charlie Turner, et al. Effectiveness of one dose of mva–bn smallpox vaccine against mpox in england using the case-coverage method: an observational study. The Lancet Infectious Diseases, 23(7):828–835, 2023.

[70] Alexandra F Dalton. Estimated effectiveness of jynneos vaccine in preventing mpox: a multijurisdictional case-control study—united states, august 19, 2022–march 31, 2023. MMWR. Morbidity and mortality weekly report, 72, 2023.

[71] Nicholas P Deputy, Joseph Deckert, Anna N Chard, Neil Sandberg, Danielle L Moulia, Eric Barkley, Alexandra F Dalton, Cory Sweet, Amanda C Cohn, David R Little, et al. Vaccine effectiveness of jynneos against mpox disease in the united states. New England Journal of Medicine, 388(26):2434–2443, 2023.

[72] Amanda B Payne. Reduced risk for mpox after receipt of 1 or 2 doses of jynneos vaccine compared with risk among unvaccinated persons—43 us jurisdictions, july 31–october 1, 2022. MMWR. Morbidity and Mortality Weekly Report, 71, 2022.

[73] Eli S Rosenberg. Effectiveness of jynneos vaccine against diagnosed mpox infection—new york, 2022. MMWR. Morbidity and mortality weekly report, 72, 2023.

[74] Yael Wolff Sagy, Roy Zucker, Ariel Hammerman, Hila Markovits, Noa Gur Arieh, Wiessam Abu Ahmad, Erez Battat, Noga Ramot, Guy Carmeli, Avner Mark-Amir, et al. Real-world effectiveness of a single dose of mpox vaccine in males. Nature Medicine, 29(3):748–752, 2023.

[75] Mikael Sunnåker, Alberto Giovanni Busetto, Elina Numminen, Jukka Corander, Matthieu Foll, and Christophe Dessimoz. Approximate bayesian computation. PLOS Computational Biology, 9(1):1–10, 01 2013.

[76] Nextstrain team. Nextstrain build for mpox virus. https://github.com/nextstrain/mpox, 2026. Commit 9128c1d, accessed 2026-05-26.

[77] Ivan Aksamentov, Cornelius Roemer, Emma B. Hodcroft, and Richard A. Neher. Nextclade: clade assignment, mutation calling and quality control for viral genomes. Journal of Open Source Software, 6(67):3773, 2021.

[78] Loris Bennett, Bernd Melchers, and Boris Proppe. Curta: A Generalpurpose High-Performance Computer at ZEDAT, Freie Universität Berlin. 10.17169/refubium-26754, 2020.

